# Resolving phenotypic and prognostic differences in interstitial lung disease related to systemic sclerosis by computed tomography-based radiomics

**DOI:** 10.1101/2020.06.09.20124800

**Authors:** J. Schniering, M. Maciukiewicz, H. S. Gabrys, M. Brunner, C. Blüthgen, C. Meier, S. Braga-Lagache, A. Uldry, M. Heller, O. Distler, M. Guckenberger, H. Fretheim, A. Hoffmann-Vold, C. T. Nakas, T. Frauenfelder, S. Tanadini-Lang, B. Maurer

## Abstract

Radiomic features are quantitative data calculated from routine medical images and have shown great potential for disease phenotyping and risk stratification in cancer. Patients with systemic sclerosis (SSc), a multi-organ autoimmune disorder, have a similarly poor prognosis (10-year survival of 66%), due to interstitial lung disease (ILD) as the primary cause of death. Here, we present the analysis of 1,355 stable radiomic features extracted from computed tomography scans from 156 SSc-ILD patients, which describe distinct disease phenotypes and show prognostic power in two independent cohorts. We derive and externally validate a first quantitative radiomic risk score, qRISSc that accurately predicts progression-free survival in SSc-ILD and outperforms current clinical stratification measures. Correlation analysis with lung proteomics, histology and gene expression data in a cross-species approach demonstrates that qRISSc reverse translates into mice and captures the fibrotic remodeling process in experimental ILD.

## Introduction

“Radiomics” is a field of research which describes the in-depth analysis of tissue phenotypes by automated and computational retrieval of high-dimensional quantitative imaging features from medical images^1,2^. This approach challenges the current practice of subjective and mostly qualitative visual image analysis and extends it beyond traditional image semantics^3^. The value of these quantitative radiomic features lies in their ability to capture comprehensive, imaging-based characteristics, including tissue intensity, shape, and texture. These features mostly cannot be perceived by eye, and, as such, contain distinct and complementary information compared to clinical, laboratory, or functional data^3^. By integrating with appropriate data mining techniques, radiomics has demonstrated great potential for evidence-based decision support in personalized medicine^1,2^. In different types of cancer, specific radiomic features have emerged as promising prognosticators for clinical outcome and drug response^4–8^. Radiomic features have further been shown to reflect the underlying tumor biology as assessed by correlation with tissue-based genomic or proteomic data^6,8–10^, supporting the hypothesis that macroscopic radiomic features allow tracing back to microscopic tissue characteristics^1^. While radiomics research has been initiated and is most advanced in oncology, including lung cancer^4,6,9,10^, its potential has not yet been explored in-depth for non-malignant lung diseases^11–15^, which account for one out of six deaths worldwide and are the third leading cause of mortality^16^.

Interstitial lung disease (ILD) is an umbrella term for a group of chronic parenchymal lung disorders with different etiologies, in which fibrosis of the lung interstitium is the common pathophysiological end-stage. Idiopathic pulmonary fibrosis (IPF) and ILD associated with systemic sclerosis (SSc) are the most prevalent and severe subtypes^17,18^. SSc is a rare autoimmune connective tissue disease with multi-organ involvement and a global incidence of 10–50 cases per million people per year^19^. ILD, which can develop early in the disease course^20^, is the leading cause of death accounting for 35% of disease-specific mortality^21,22^. A major challenge in the management of SSc-ILD patients is the large inter-individual heterogeneity on a clinical and molecular level^23–25^. Whereas approximately 46% of SSc-ILD patients will develop progressive disease with need of treatment, others will remain stable for years even without therapeutic intervention^26^. In addition, the progression rate i.e. time to and pace of progression, varies tremendously^26^. This high variability in patient-specific disease trajectories warrants valid prognostic biomarkers for individual risk stratification.

Medical imaging, particularly high resolution computed tomography (HRCT), is an integral part of the standard-of-care of SSc-ILD patients, as it is non-invasive and allows longitudinal monitoring of the entire lung pathology with high sensitivity^27–31^. Pulmonary function tests (PFTs)^32^ are another commonly used diagnostic tool, although they lack disease specificity and fail as a stand-alone screening method in early SSc-ILD with preserved lung volumes despite radiologically proven parenchymal disease^33–35^. Extent of lung fibrosis on HRCT (threshold of 20%) and/or presence of a restrictive ventilation defect (Forced Vital Capacity (FVC)% predicted < 70%) have been associated with poor SSc-ILD outcome^36,37^ and are currently most often used for staging and risk stratification in clinical practice. Recent data challenge this convention demonstrating that even patients with mild SSc-ILD (lung fibrosis extent on HRCT <10%, FVC 80-100%) frequently developed progressive ILD with reduced survival^26^.

The lack of prognostic and independently validated biomarkers^38,39^, which could be met by the recent advances in image acquisition, processing, and high-throughput image analysis, have prompted the herein presented study.

Here, we report four key findings: 1) We confirm the reproducibility of radiomic features with respect to tissue segmentation and identify 1,355 stable radiomic features for SSc-ILD. 2) By using unsupervised clustering, we discover two distinct patient clusters based on radiomic profiling that describe different SSc-ILD phenotypes and provide crucial information on differences in (progression-free) survival. 3) We derive and validate a prognostic radiomic risk score for SSc-ILD progression, qRISSc, that allows accurate prediction of progression-free survival outperforming current SSc-ILD risk stratification tools in two independent, well-characterized patient cohorts. Lastly, we provide insights into the biological basis of qRISSc by correlation analysis with molecular data including whole-lung tissue proteomics, histological and gene expression data in a cross-species approach, which shows that qRISSc is reverse translatable to mice and specifically reflects the underlying fibrotic remodeling processes in (experimental) ILD.

## Results

### Study Design and Datasets

In this study, we retrospectively investigated two independent prospective cohorts of SSc-ILD patients including 90 patients (76.7% female, median age 57.5 years) from the University Hospital Zurich and 66 patients (75.8% female, median age 61.0 years) from the Oslo University Hospital. A third dataset, derived from an experimental cohort of 30 mice with bleomycin-induced lung fibrosis, a widely acknowledged preclinical model for ILD^40^, was used for association studies with biological features, including proteomic, histological, and gene expression data. For every subject, we defined and extracted 1,386 radiomic features (Supplementary File 1) from CT images including 17 intensity, 137 texture, and 1,232 wavelet features using our in-house developed radiomics software Z-Rad. A detailed description of the study workflow is available in Figure 1. A summary of patients’ demographics and clinical characteristics at baseline for both study cohorts is given in Supplementary Table 1.

**Figure 1:**
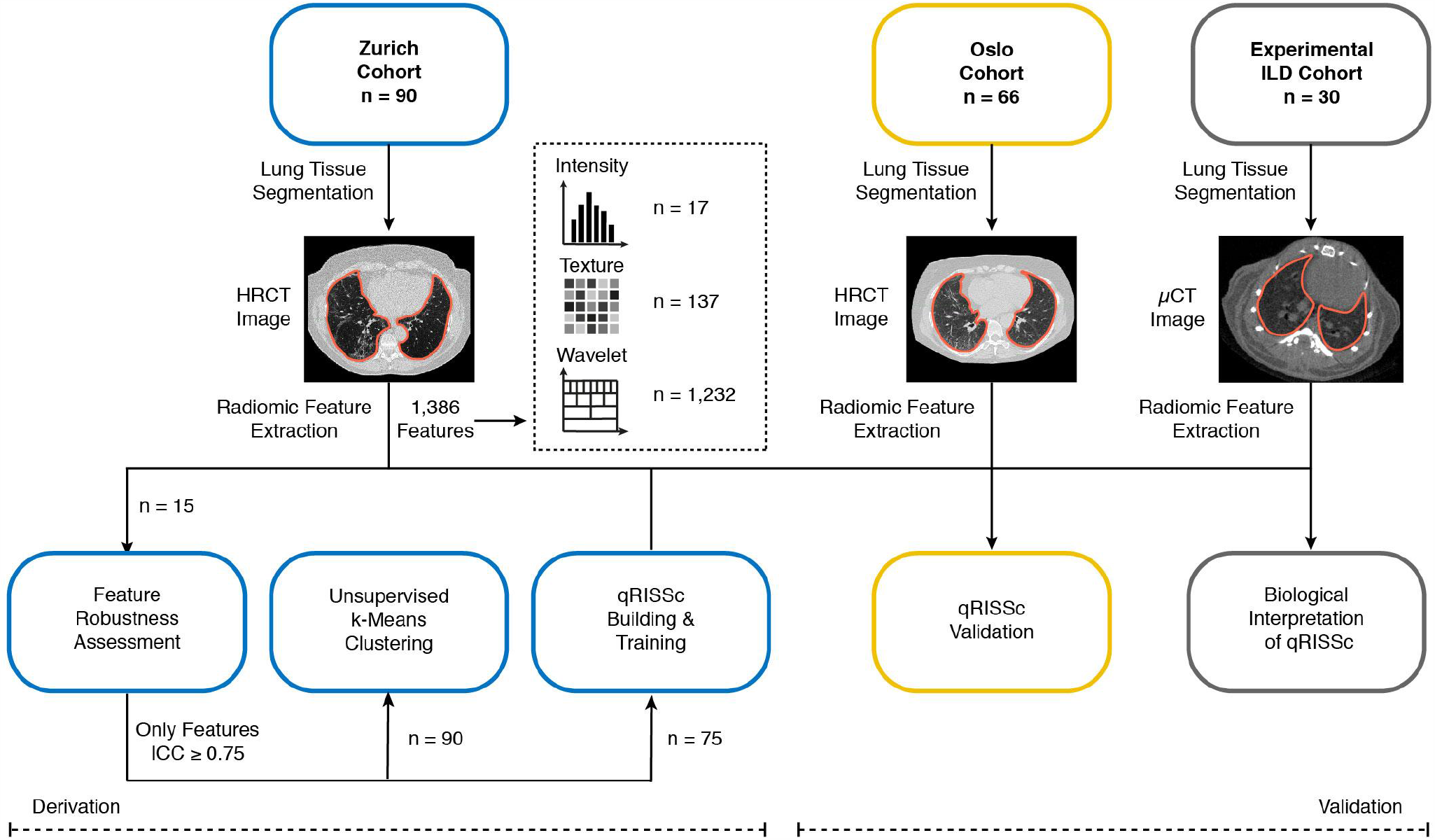
Study workflow. Our in-house developed radiomics software Z-Rad was applied to three different datasets, including two independent cohorts of SSc-ILD patients from 1) the University Hospital Zurich (derivation cohort) and 2) the Oslo University Hospital (validation cohort), and one experimental ILD cohort, composed of 30 bleomycin-treated mice for association studies with biological features (i.e. proteomic, histological, and gene expression data). For every subject, in total, 1,386 radiomic features were extracted from segmented CT images, including 17 intensity, 137 texture, and 1,232 wavelet features. Filtering of robust radiomic features (ICC ≥ 0.75), unsupervised clustering, and construction of the quantitative radiomic ILD risk score (qRISSc) for progression-free survival in SSc-ILD were performed in the Zurich cohort. Independent and external validation of the built qRISSc was performed using the Oslo cohort.

### Identification of robust radiomic features in SSc-ILD

Stability and reproducibility of radiomic features is a prerequisite for building generally applicable prognostic and predictive radiomic models^41^. Operator variability in the tissue segmentation process can largely affect radiomic feature values by introducing errors in the calculation of features and can thus impact prediction accuracy^6,42,43^. In the context of SSc-ILD, robustness of radiomic features against semi-automated lung delineation has not yet been evaluated. Hence, we first assessed the intra- and inter-operator feature stability by intraclass correlation (ICC) analysis in a subgroup of SSc-ILD patients from the Zurich cohort (*n* = 15, Fig. 2).

**Figure 2:**
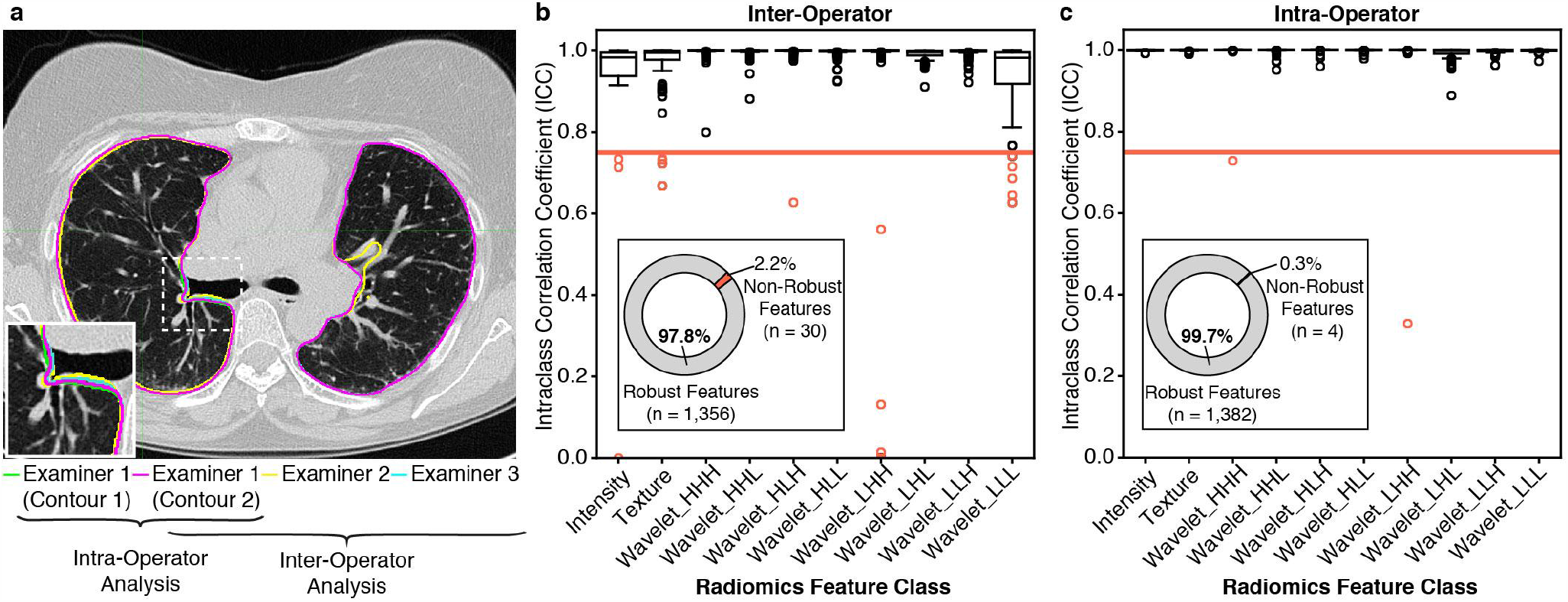
Assessment of radiomic feature robustness against inter-and intra-operator variability in the semi-automated lung segmentation process. (**a**) Representative transversal HRCT image showing the lung delineation structures of the three different examiners (examiner 1: green and magenta, examiner 2: yellow, examiner 3: cyan) for the intra- and inter-operator ICC analyses. (**b**) Boxplots showing the distribution of the ICC coefficient per radiomic feature category for inter-operator ICC analysis and (**c**) intra-operator ICC analysis. In (**b, c**), the bright red line indicates the threshold defined for the ICC analyses (ICC = 0.75; corresponding to good reproducibility^44^). The pie charts summarize the respective percentage and total numbers of robust (ICC ≥ 0.75) and non-robust (ICC < 0.75) radiomic features.

Overall, excellent agreement and overlap in the semi-automatically delineated lung structures between the different operators was observed (Fig. 2a), confirming the reproducibility and validity of our lung segmentation protocol. Accordingly, the vast majority of radiomic features (97.8%) was found to be robust with only a few features having lower ICC values than 0.75^44^ in the inter-operator (2.2%, *n* = 30, Fig. 2b) and intra-operator (0.3%, *n* = 4, Fig. 2c) analysis resulting in a final set of 1,355 robust radiomic features for SSc-ILD.

### Unsupervised clustering of radiomic features identified two distinct SSc-ILD patient clusters

Having confirmed the robustness of radiomic features in SSc-ILD, we next explored the radiomic phenotypes of the 90 SSc-ILD patients from the Zurich cohort with unsupervised *k*-Means clustering in order to identify homogeneous imaging-based groups without any a priori assumption. We then examined their associations with clinical baseline parameters and patient outcome among the obtained clusters.

Using the final set of 1,355 robust features, cluster analysis revealed two distinct patient clusters based on their radiomic profiles with good bootstrap (*n* = 1,000) cluster stability (Jaccard coefficient for cluster 1: 0.90 and for cluster 2: 0.82, wherein 1 indicates perfect stability; Fig. 3a and b). The two clusters exhibited different baseline characteristics (Fig. 3 and Supplementary Tab. 2) with patients from cluster 2 (*n* = 31) having a significantly worse restrictive ventilation defect than patients in cluster 1 (*n* = 59) as evidenced by significantly lower lung function parameters (*p* < 0.001, Fig. 3c to e). In addition, patients from cluster 2 performed significantly worse in the 6-min walk test, a measure of the exercise capacity of the patient, and showed lower oxygen saturation at the beginning and end of the test, shorter walk distance, and more severe exertion (Fig. 3g to j). There was also a significant association of cluster 2 with the presence of pulmonary hypertension (*p* = 0.001, Fig. 3a), which was consistent with the significantly higher mean systolic pulmonary artery pressure (PAPsys; measured by right heart catheterization and/or echocardiography) in subjects belonging to this cluster (Fig. 3f). Cluster 2 was further significantly enriched for certain visual HRCT patterns such as honeycombing (*p* = 0.004) and the radiological usual interstitial pneumonia (UIP) subtype (*p* = 0.01, Fig. 3a). Yet, the clusters were not found to be associated with the different radiomic feature classes (Fig. 3a).

**Figure 3:**
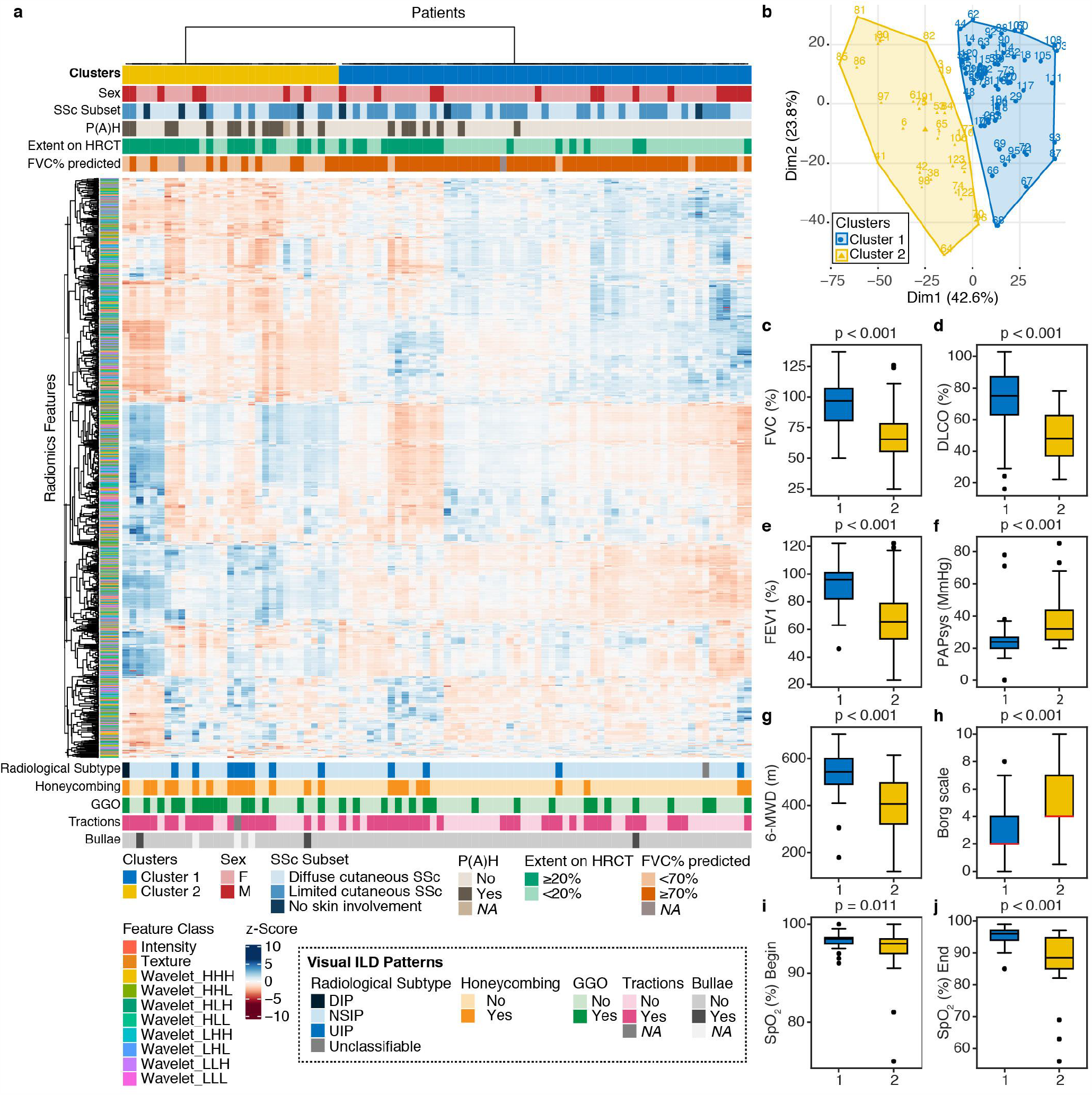
Unsupervised *k*-Means clustering of radiomic data from SSc-ILD patients. (**a**) Heatmap summarizing the *k*-Means clustering results (Zurich cohort, *n* = 90). Prior to clustering, radiomic features were z-scored. Associations between the two identified radiomic clusters with baseline clinical parameters (above) and visual ILD patterns depicted on HRCT (below) are shown. (**b**) *k*-Means cluster plot indicating two stable clusters (cluster 1: 0.90, in blue and cluster 2: 0.82, in yellow). (**c-e**) Boxplots comparing baseline lung function parameters between the two clusters, including FVC% predicted (**c**), DLCO% predicted (**d**) and FEV1% predicted (**e**). Boxplot showing the distribution of the systolic pulmonary artery pressure (PAPsys) per cluster. (**g-j**) Boxplots indicating the results from the 6-min walk test, including 6-min walk distance (6-MWD, **g**), Borg scale of perceived exertion (**h**, scale 0-10, 0 = no exertion, 1 = very weak, 2 = weak, 3 = moderate, 5 = strong, 7 = very strong, 10 = extreme exertion). Abbreviations: SSc = systemic sclerosis, P(A)H = pulmonary (arterial) hypertension, FVC = forced vital capacity, FEV1 = forced expiratory volume in 1 second, DLCO = diffusing capacity for carbon monoxide, F = female, M = male, DIP = diffuse interstitial pneumonia, NSIP = nonspecific interstitial pneumonia, UIP = usual interstitial pneumonia, GGO = ground glass opacification.

Most notably, radiomics clusters did not stratify patients according to classical definitions of ILD severity, including limited and extensive disease extent as defined by HRCT analysis (<20%, or ≥20%) or PFTs (FVC% predicted ≥70% or <70%), respectively, although significant associations with both disease classifiers were detected (*p* = 0.002 and *p* < 0.001, respectively). Furthermore, there was no difference in general disease characteristics, including age, gender distribution, SSc disease duration, active immunomodulatory therapy, extent of skin involvement, and autoantibody profiles between the two clusters (Fig. 3a and Supplementary Tab. 2), suggesting that radiomic profiles capture lung disease-specific phenotypic differences among SSc-ILD patients.

We then assessed whether the two identified patient clusters also differed in terms of their clinical outcome by survival analysis with Kaplan-Meier estimator. Consistent with their worse disease phenotype, patients of cluster 2 also showed a higher probability of faster disease progression and a decrease in progression-free survival defined by either the time to a relative decline of ≥ 15% in FVC% predicted (*p* = 0.001, HR = 3.52, 95% CI = (1.66-7.45), Fig. 4a) or the time to decline assessed by a recently proposed FVC-DLCO composite index^45^ (*p* = 0.005, HR = 2.73, 95% CI = (1.36 - 5.50), Fig. 4b). In addition, a marginal association with overall survival was detected, suggesting a higher risk of all-cause death for patients of cluster 2 (*p* = 0.07, HR = 2.37, 95% CI = (0.97 - 5.98), Fig. 4c). This demonstrates that radiomic features do not only capture ILD-specific phenotypic differences but also contain important prognostic information. In summary, unsupervised clustering of the radiomic profiles distinguished and characterized distinct SSc-ILD phenotypes and provided crucial information on differences in (progression-free) survival.

**Figure 4:**
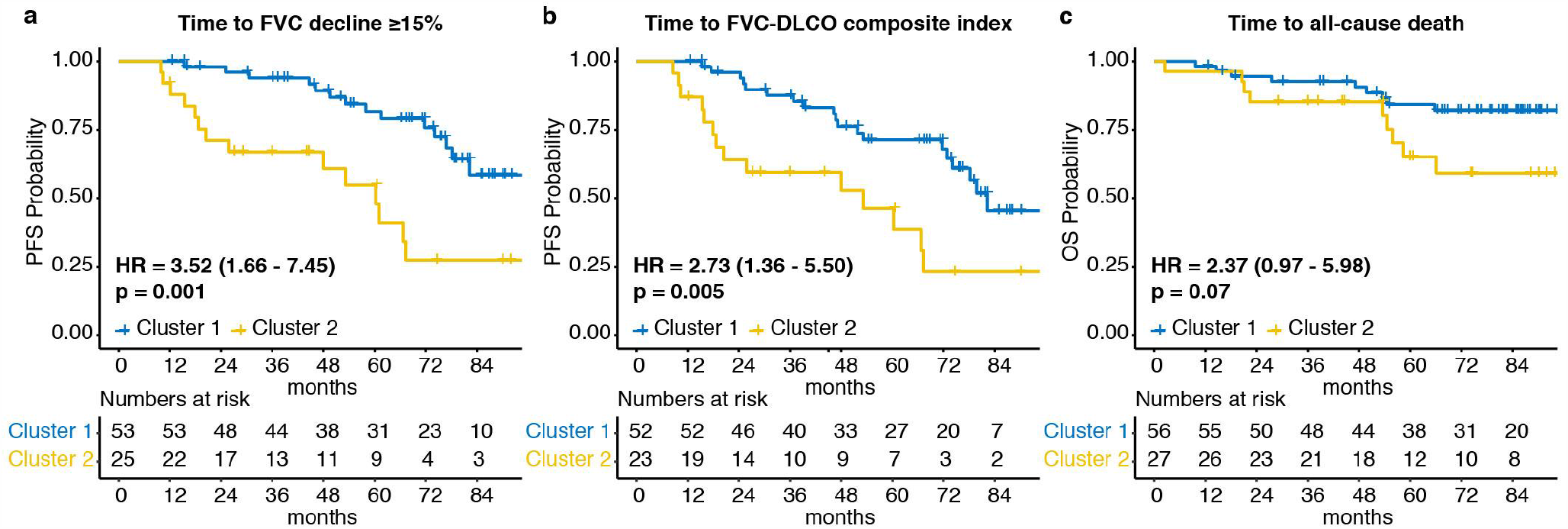
Associations of radiomic clusters with clinical outcome. (**a, b**) Kaplan Meier curves for progression-free survival (PFS) defined as either the time to relative FVC decline ≥ 15% (**a**) or the time to the FVC-DLCO composite index (**b**, FVC-DLCO composite index = relative decrease in FVC% predicted of ≥15%, or a relative decline in FVC% predicted of ≥10% combined with DLCO% predicted of ≥15% according to ^45^). (**c**) Kaplan Meier curves for overall survival (OS) defined as the time to all-cause death. The Hazard ratios (HR) with 95% confidence intervals and *p* value of the univariate Cox regression are shown.

### Quantitative radiomic risk score predicted progression-free survival in SSc-ILD

The high inter-individual variability in the clinical course of SSc-ILD warrants valid prognostic biomarkers for personalized management, which so far are lacking. Tools to guide treatment decisions, i.e. watch & wait in stable ILD versus immunomodulation in progressive ILD, based on individual risk stratification would have tremendous clinical impact. Having found that radiomic features are able to distinguish between “high” and “low risk” clusters of SSc-ILD patients for ILD progression, we next assessed the potential to build prognostic radiomic signatures allowing risk stratification without the need of extracting all radiomic features.

To this end, we derived a quantitative radiomics score following a similar design as recently proposed by Lu et al.^8^ for risk stratification in ovarian cancer. The Zurich cohort was used for score construction and independent validation was performed on an external out-of-sample dataset consisting of 66 SSc-ILD patients from Oslo. Following the approach by Lu et al.^8^, we first removed radiomic features without predictive power using univariate Cox regression analyses for progression-free survival based on our primary study endpoint defined as the time to a relative FVC% decline ≥ 15%. With this approach, in total, 32 prognostic radiomic features were retained. In a second step, we applied cross-validated LASSO penalized regression to the retained features and selected all features with a non-zero LASSO coefficient. This resulted in *j* = 26 features, including four intensity, nine texture and 13 wavelet-filtered features. To construct the final quantitative radiomic risk score for progression-free survival in SSc-ILD (qRISSc), we calculated the weighted sum of the selected radiomic features, 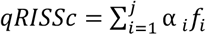 with α being the feature weight and *f* being the values of z-transformed radiomic features. Due to the rather low sample size of our derivation cohort (*n* = 75), which is within the expected range for this rare disease, we did not perform weighting of score features by LASSO coefficient as originally done by Lu et al.^8^ to avoid overestimation of the effect sizes. Instead, we followed a maximum-likelihood approach assigning equal feature weights of 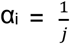 to calculate qRISSc. Based on the Zurich cohort, we determined an optimal cut-off for qRISSc (cut-off,value = 0.21) that best split SSc-ILD patients into high- and low-risk groups, with high-risk patients having a higher probability of earlier lung function decline than low-risk patients (median progression-free survival time = 48.0 months vs. 82.30 months; Fig. 5a).

**Figure 5:**
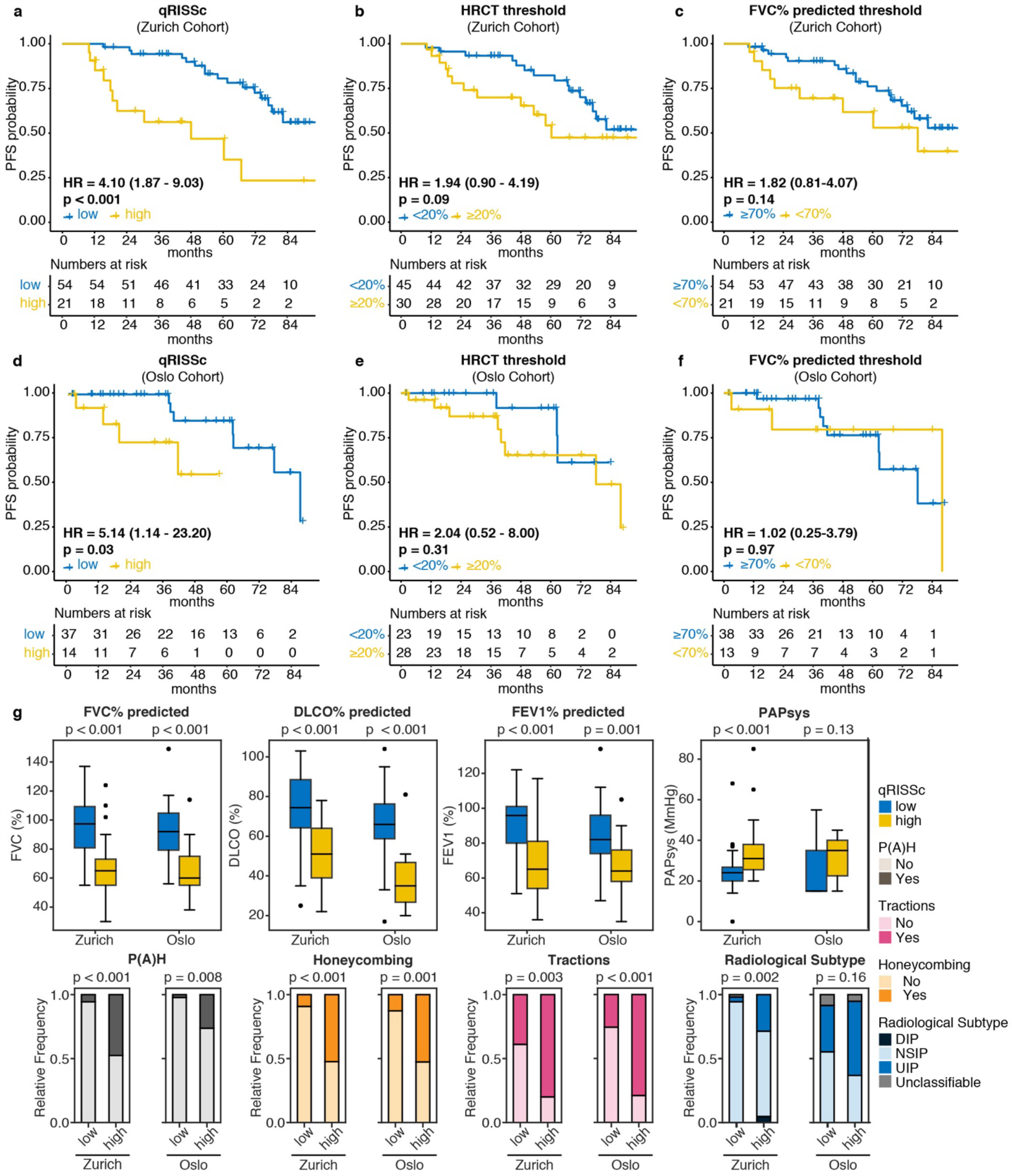
Prognostic performance of qRISSc compared to standard SSc-ILD staging systems, and associations of qRISSc with clinical parameters at baseline. **(a-c)** Kaplan Meier curves of (**a**) the constructed quantitative radiomic ILD risk score (qRISSc), (**b**) the HRCT threshold and (**c**) the FVC% predicted threshold for progression-free survival (PFS) defined as the time to relative FVC decline ≥ 15% in the derivation (Zurich) cohort (*n* = 75). (**d-f)** Kaplan Meier curves of (**d**) the constructed quantitative radiomic ILD risk score (qRISSc), (**e**) the HRCT threshold and (**f**) the FVC% predicted threshold for progression-free survival (PFS) in the validation (Oslo) cohort (*n* = 51 with complete survival information). (**g**) Significant associations of qRISSc with clinical baseline parameters in both, the derivation (Zurich) cohort (*n* = 75) and validation (Oslo) cohort (*n* = 66). For (**a-f**), the Hazard ratios (HR) with 95% confidence intervals and *p* value of the univariate Cox regression are shown. For (**g**), Fisher’s exact test was used for comparison of categorical and Mann-Whitney U for comparison of numerical variables, respectively.

To validate the score, we then independently applied qRISSc using the same cut-off value on the external out-of-sample dataset from Oslo. Notably, in this independent dataset, patients stratified by qRISSc were also significantly different in their progression-free survival with a median progression-free survival time of 41.7 months in the high-risk versus 88 months in the low-risk group (*p* = 0.03, HR = 5.14, 95% CI = (1.14 - 23.20) Fig. 5d), thus validating the score.

We next compared the prognostic potential of qRISSc to existing clinical SSc-ILD stratification tools, including subgrouping of patients based on disease extent on HRCT (<20% or ≥20% fibrosis) or the FVC% predicted threshold of <70%, respectively^35^. In both the external validation cohort from Oslo and the derivation cohort from Zurich, neither fibrosis extent on HRCT nor the FVC%-based patients’ risk stratification was prognostic for future lung function decline in univariate analysis, overall indicating the superiority of qRISSc over current state-of-the art prognostic measures (Fig. 5b and c and 5e and f).

To evaluate the prognostic power of qRISSc, we calculated the concordance index (C-index) for the survival analysis, which is the equivalent to the area under the receiver operating characteristic curve^46^. In both the training and validation cohort, qRISSc showed good prognostic performance with a C-index of 0.67 (standard error = 0.05; *p* < 0.001) and 0.71 (standard error = 0.07; *p* = 0.03), respectively. Of note, in both study cohorts, qRISSc was also marginally associated with the other two clinical outcome parameters, including the FVC-DLCO composite index (Zurich cohort: *p* = 0.002, Oslo cohort: *p* = 0.06) and overall survival (Zurich cohort: *p* = 0.001, Oslo cohort: *p* = 0.11).

qRISSc-stratified high- and low-risk patients’ groups also revealed distinct differences in their clinical baseline parameters (Fig. 5g, Supplementary Tabs. 4 and 5). High-risk patients consistently presented with a more severe ILD phenotype with worse lung function parameters at baseline compared to low risk patients in both study cohorts (Fig. 5g). A high-risk score was further associated with the presence of pulmonary hypertension, the extent of fibrosis on HRCT (Supplementary Tabs. 4 and 5), and certain visual ILD HRCT patterns including honeycombing, traction bronchiectasis and UIP radiological subtype (Fig. 5g). Interestingly, no differences were observed in general SSc disease characteristics and patient demographics including age, sex, disease duration, and extent of skin involvement (Supplementary Tabs. 4 and 5).

Since radiomic features are greatly affected by image acquisition and reconstruction protocols^41^, we also assessed the influence of the different scan parameters, including convolution kernel, slice thickness, and type of CT scanner used in this study (Supplementary Tab. 6) on the distribution of qRISSc. Multidimensional scaling (MDS) of combined SSc-ILD patients from Zurich and Oslo cohort revealed that qRISSc did not separate groups according to different imaging sites and scan parameters, including differences in slice thickness (range 0.6 to 3 mm), reconstruction kernels, and scanner types (Supplementary Fig. 1).

In conclusion, we derived and validated a quantitative radiomic risk score for SSc-ILD progression, qRISSc, which accurately predicted progression-free survival and outperformed currently existing prognostic measures in an out-of-sample, external patient cohort, confirming the prognostic value of radiomic features and highlighting the potential of radiomic feature-based clinical scores for risk stratification in SSc-ILD.

### Defining the pathophysiological basis of the quantitative radiomic risk score

Finally, we aimed to define the biological basis and to reveal possible associations of qRISSc with the underlying ILD pathophysiology. Current pathogenic models suggest that SSc-ILD is triggered by repeated epithelial and microvascular injury initiating inflammation with activation of innate and adaptive immune responses, which results in myofibroblast activation and increased deposition of extracellular matrix (ECM) ultimately causing the disruption of the lung architecture, functional impairment and even respiratory failure^47^.

Lung biopsies are only very rarely performed in SSc-ILD patients^48^ and patient-derived tissue samples for molecular analysis are scarce. Since, consequently, imaging-matched human biosamples were not available for this study, we conducted a cross-species correlation approach, using the mouse model of bleomycin-induced lung fibrosis as a model system for SSc-ILD. Bleomycin-induced lung fibrosis is the most extensively used and best characterized preclinical animal model for ILD as it resembles many aspects of the human ILD pathophysiology including epithelial cell apoptosis, inflammatory cell infiltrates, and extracellular matrix (ECM) deposition^49,50^. In addition, we have recently confirmed that also radiomic signatures largely translate between experimental ILD in bleomycin-treated mice and ILD in SSc patients^51^. Analogously to SSc-ILD patients, we extracted the complete set of radiomic features from segmented micro-CT images of bleomycin-treated mice (*n* = 30), applied z-transformation and built the equally weighted sum of the 26 previously selected radiomic score features to construct qRISSc in mice. We firstly compared the quantitative score values obtained in mice and our two patients’ cohorts to ensure that qRISSc is (reverse) translatable from SSc-ILD patients to mice. We found a very similar score distribution between all three datasets derived from mice and the two patient cohorts, confirming the suitability of this animal model as a preclinical “radiomic surrogate” for human ILD (Fig. 6a).

**Figure 6.**
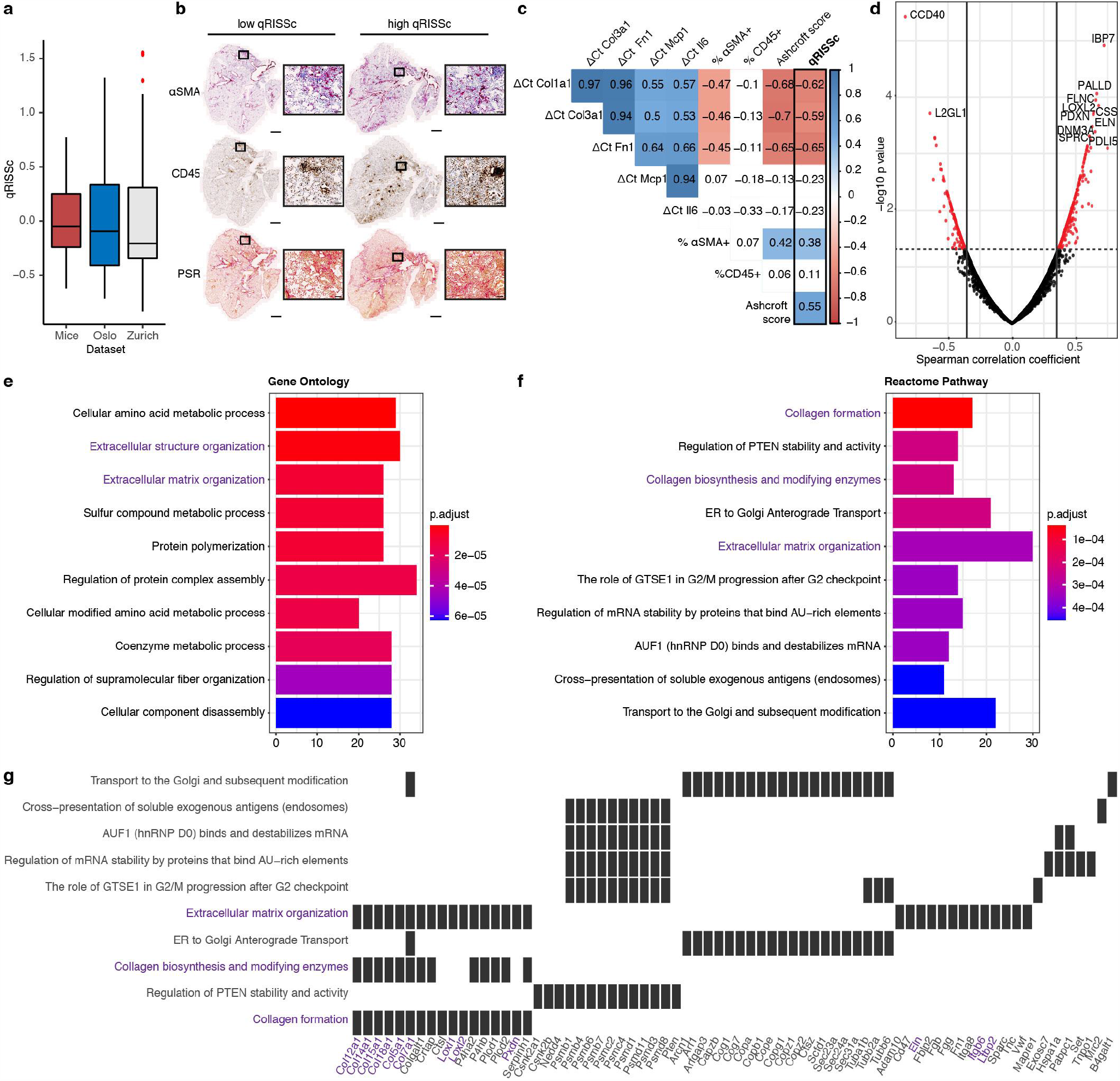
Correlation analysis of qRISSc with molecular data in experimental ILD. (**a**) Score distribution across the three datasets, demonstrating a similar qRISSc distribution between mice of the bleomycin-induced lung fibrosis model (*n* = 30) and SSc-ILD patients (Zurich, *n* = 75; Oslo, *n* = 66). (**b**) Representative histological images of bleomycin-treated mice with low and high qRISSc that were stained for the myofibroblast marker alpha smooth muscle actin (αSMA, upper panel), the pan-leukocyte marker CD45 (middle panel) and picrosirius red to visualize collagen fibers (PSR, collagen = red, lower panel). Sections of the entire right caudal lobe (scale bar = 1 mm) with higher magnification views (100x magnification, scale bar = 100 μm) are shown. (**c**) Correlation matrix for qRISSc with histological parameters (percentage of αSMA and CD45 positivity, and Ashcroft score), and messenger RNA (mRNA) expression of inflammatory (*Il6, Mcp1*) and fibrotic (*Col1a1, Col3a1, Fn1*) genes. A lower ΔCt value and thus negative correlation indicates higher gene expression. The Spearman correlation coefficient *rho* is shown. Non-significant associations are depicted in white. (**d**) Volcano plot for qRISSc-correlated proteins. Proteins with *rho* ≥ |0.3| and *p* < 0.05 are highlighted in red. (**e**) Bar plot of the top 10 (based on p-value) biological processes associated with qRISSc. (**f**) Bar plot of the top 10 (based on p-value) pathways associated with qRISSc. (**g**) Heatplot indicating the top enriched proteins per molecular pathway. For (**e-g**), the most important associations are highlighted in purple. For pathway analyses, only proteins with *rho* ≥ |0.3| and *p* < 0.05 were considered.

Next, we performed pathway enrichment analysis for significantly qRISSc-correlated proteins (*rho* ≥ |0.3|, p<0.05) derived from whole-lung tissue proteomics to reveal associations of qRISSc with molecular pathways and processes related to ILD using Reactome^52^ and GO:Biological Process^53^ databases. In total, we found 634 out of 5,311 identified proteins (11.94%) that significantly correlated with qRISSc (Fig. 6d). Pathway enrichment analysis of those proteins revealed that pathways related to fibrosis development, particularly pathways associated with ECM organization and formation were most significantly associated with qRISSc (Fig. 6f and g). Consistently, the enriched biological processes that significantly correlated with qRISSc were also largely linked to pro-fibrotic remodeling processes underlying ILD, including processes related to protein polymerization and ECM assembly (Fig. 6e).

Among the highly and significantly qRISSc-correlated proteins were multiple ECM proteins, such as collagen 5α1, (CO5A1, *rho* = 0.48), collagen 7α1 (CO7A1, *rho =* 0.55), collagen 12α1 (COCA1, *rho =* 0.46), collagen 15α1 (COFA1, *rho* = 0.48), collagen 18α1 (COIA1, *rho* = 0.47), filamin-C (FLNC, *rho* = 0.66), and elastin (ELN, *rho* = 0.63) as well as proteins required for ECM assembly and crosslinking, including members of the lysyl oxidase family, such as LOXL1 (*rho* = 0.56) and LOXL2 (*rho* = 0.68), or peroxidasin (PXDN, *rho* = 0.64). In addition, proteins involved in TGF-β activation, including latent-transforming growth factor beta-binding protein 2 (LTBP2: *rho* = 0.50) and integrin β6 (ITB6, *rho =* 0.55), were strongly correlated with qRISSc (Fig. 6g).

To complement the proteomic analysis, we additionally performed whole-slide digital histopathological and gene expression analysis of established major fibrotic and inflammatory markers^50,54,55^ (Fig. 6b and c). In line with the proteomic data, qRISSc was also significantly correlated with fibrotic markers on histological level with a higher qRISSc value corresponding to a higher fibrosis score (Ashcroft score^56^, *rho* = 0.55), and increased expression of αSMA, a marker for activated fibroblasts (*rho* = 0.38). Consistently, qRISSc also showed significant association with the expression of fibrotic genes, including collagen 1α1 (*Col1a1, rho =* -0.62), collagen 3α1 (*Col3a1, rho =* -0.59), and fibronectin 1 (*Fn1, rho =* -0.65), where a lower ΔCt value and thus negative correlation indicates higher gene expression. Most notably, neither on histological nor on gene level, qRISSc correlated with inflammatory markers, such as the number of CD45+ inflammatory cells in tissue sections, interleukin 6 (*Il6*), and monocyte chemoattractant protein 1 (*Mcp1*) mRNA expression (Fig. 6b and c). Collectively, this suggests that qRISSc specifically reflects the underlying fibrotic remodeling processes in (experimental) ILD. The fact that we observed a similar score distribution between experimental and human ILD demonstrated that qRISSc was reverse translatable from humans to mice, pointing out the potential value of the bleomycin-induced lung fibrosis model as a preclinical “radiomic surrogate” for human ILD.

## Discussion

As the major cause of morbidity and mortality in SSc, ILD remains a challenge for researchers and physicians although targeted therapies are becoming increasingly available^57^. The lack of validated prognostic biomarkers^39^ opposes individualized patient management. Traditionally, molecular data derived from tissue biopsies, including genomic and proteomic data have been explored for precision medicine approaches. However, the invasiveness of tissue biopsies, the unsuitability for longitudinal assessments, the high risk of non-representative sampling due to spatial disease heterogeneity and also the high costs associated with molecular profiling have largely limited the implementation in clinical practice. This applies even more to SSc-ILD, where lung biopsies are only conducted in exceptional cases, and are not required for establishing the diagnosis^48^. With our radiomics study, we aimed to advance the development of personalized medicine approaches in SSc-ILD by providing a non-invasive, cost-effective and reproducible way of evaluating phenotypic information derived from HRCT as a base for individualized risk stratification. In this study, we identified and analyzed 1,355 stable and reproducible HRCT-derived radiomic features from 156 SSc-ILD patients. Using unsupervised clustering and supervised prediction modelling we showed that radiomic profiles can quantify lung-specific phenotypic differences based on image intensity, texture, and wavelet transformation and that they have prognostic power. We further derived and validated a first quantitative radiomic risk score for SSc-ILD progression, qRISSc, that adequately predicted progression-free survival in two independent cohorts. For this, we adapted the recently proposed methodology for radiomics-based score design from Lu et al.^8^, which was originally developed for risk stratification and drug response prediction in epithelial ovarian cancer. The same methodology for feature selection and score building was very recently also adapted by Vaidya et al. and successfully applied to non-small cell lung cancer for prediction of added benefit from adjuvant chemotherapy and disease-free survival^58^. Herein, we demonstrated that this approach is also applicable outside of the cancer field and can be transferred to a non-malignant lung disease emphasizing the general applicability and great value of radiomics and radiomics-derived scores to support and complement clinical decision making.

In both study cohorts, radiomics-based high-risk stratification was consistently associated with a more severe ILD phenotype at baseline, presence of pulmonary hypertension and certain visual ILD HRCT patterns including honeycombing, traction bronchiectasis and UIP radiological subtype, all of which have been discussed as potential risk factors in SSc-ILD^39,59^. The fact that we did not observe correlations with other suggested clinical risk factors such as e.g. diffuse cutaneous SSc subset, older age, male sex, and anti-topoisomerase 1-positivity^32,60^, underlines that radiomic features capture lung-specific information independent of demographic and clinicoserological characteristics. Proposed prognostic factors and/or models for SSc-ILD vary among studies and patient cohorts^39,59^. At present, disease extent on HRCT and FVC <70% at first presentation are the most commonly used markers for risk stratification and guidance of treatment decision in clinical practice^35–37^. Our study showed that both stratification tools failed to predict progression-free survival in two independent cohorts. The superior performance of radiomics might arise from the added value of more detailed and in-depth information on lung pathology. Tissue heterogeneity is reflected on different spatial levels comprising e.g. the radiological, macroscopic, cellular, and the molecular level. In “radiomic terms”, tissue heterogeneity is best described by texture features, which are built upon density-based assessments. They identify different patterns in the image by describing voxel intensities and their spatial arrangement^61^. In our study, most features in qRISSc (e.g. “coarseness”, “cluster tendency”, “sum of variance”) belonged to the radiomics class of texture features or of wavelet transformations thereof. Our results are in accordance with previous studies, where texture features seemed to outperform first order (intensity) features for prognostic purposes^6,8,10^.

The hypothesis that radiologic (radiomic) phenotype characteristics may reflect the underlying pathophysiology was supported in our study, where we used a cross-species approach integrating imaging with molecular data to define the biological basis of qRISSc. In experimental ILD, the radiomics-derived signature was closely linked to specific fibrotic remodeling processes in particular those related to ECM assembly and biosynthesis, yet did not correlate with inflammation as assessed by proteomic, histological, and immunohistochemical analyses. The fibrotic pathway activation ties in with the worse outcome of the high-risk group of SSc-ILD patients identified by qRISSc and may provide a rationale for favoring anti-fibrotic over immunomodulating or anti-inflammatory therapeutic strategies in these patients^57^. The ability of radiomic markers to specifically reflect the entire lung pathology non-invasively is particularly attractive in a complex multi-organ disease with high molecular heterogeneity such as SSc^62^. The good transferability of radiomics signatures between experimental ILD in bleomycin-treated mice and human SSc-ILD is supported by our previous study^51^ and by the similar distribution of qRISSc between all three datasets. In addition, the comparability of both imaging and molecular changes in experimental ILD induced by bleomycin with human ILD^49,51,54,55,63^ support the suitability of this animal model as preclinical “radiomic surrogate” for human ILD. The fact that radiomic features, including qRISSc, were reverse translatable from humans to mice demonstrates that well characterized and representative animal models could prove valuable to test defined hypotheses in radiomics research, particularly for studying links with pathophysiology in rare diseases with low numbers of patients and limited access to biosamples.

There are some limitations to our study, which mainly arise from its retrospective design and the rather low numbers of subjects involved. This is balanced by the high quality of the data from two independent, prospectively followed SSc cohorts from academic expert sites^64^, where patients with this rare disease are seen with regular and standardized follow-up visits. Due to the modest sample size of our derivation cohort (*n* = 75), we lacked power to assess variable importance (measured by coefficients from LASSO) and assigned equal importance to each feature following a maximum-likelihood approach to construct qRISSc. Notably, despite this fact, we could fit a significant univariate model with good prognostic power on the independent and external validation dataset, demonstrating the reproducibility and validity of the score. Future studies on larger cohorts are needed to determine feature importance and to perform proper weighting of score features, which will in turn enable us to further optimize the predictive ability of qRISSc. Concerns about the reproducibility of radiomic features arise from their dependency on image acquisition and reconstruction methodologies as well as the intra-/inter-observer variability during image segmentation^65,66^. In our study, radiomic features, including qRISSc proved to be highly stable against semi-automated lung segmentation. In addition, no batch-effects in relation to different CT scanner types, scan and reconstruction protocols across two inhomogeneous cohorts of patients from independent sites occurred. This emphasizes the translational potential of our results and is a strong argument for the future clinical application of radiomics. Since radiomic analysis can be performed on information extracted from a patient’s routine HRCT scan, radiomic scores, such as qRISSc could provide easy and readily accessible means to identify patients at higher risk of progression, who require closer monitoring and immediate therapeutic intervention.

For ultimate validation as a prognostic tool for risk stratification in SSc-ILD prospective multi-center studies and/or the analysis of retrospective randomized clinical trial datasets are required. Ideally, in these settings the performance of qRISSc in combination with suggested clinical risk factors and in comparison to other proposed clinical and functional composite scores for SSc-ILD should be assessed^45,67–69^. Within this context, the comparison of the prognostic accuracy and power of qRISSc with other HRCT-based quantitative imaging scores^27,70,71^ would also be of great interest. Ultimately, qRISSc should be validated as potential tissue surrogate by proteomic analysis of human SSc-ILD lung biopsies, which, however, is complicated by the scarce availability of tissue samples and the underlying molecular heterogeneity. To further explore the potential of qRISSc, future follow-up studies are needed, addressing its predictive power for drug response, its potential to enrich study populations for clinical trials, and most importantly, its applicability to other (fibrosing) subtypes of ILD, particularly IPF.

In summary, by integrating unsupervised cluster analysis and supervised prediction modelling, we demonstrated that radiomic features and radiomics-derived scores provide important phenotypic and prognostic information with great potential for risk stratification in SSc-ILD. In a cross-species approach, we further showed that radiomic signatures can be reverse translated from human to experimental ILD offering a valuable test system for radiomics research, which is particularly attractive for rare diseases with low numbers of patients and scarce availability of biosamples.

In conclusion, as a non-invasive, repeatable and cost-effective way of acquiring phenotypic and prognostic information derived from standard-of-care HRCT images, we provide a template approach which is potentially applicable to other forms of ILD and non-malignant lung diseases.

## Methods

### Patient Cohorts and Clinical Data

In this study, 90 patients (76.7% female, median age 57.5 years) from the University Hospital Zurich’s (derivation cohort) and 66 patients (75.8% female, median age 61.0 years) from the Oslo University Hospital’s prospective SSc patient cohorts (external validation cohort) were included. Both centers are part of the EUSTAR (European Scleroderma Trial and Research) network^72^. Patients were retrospectively selected based on the following criteria:

1. Fulfillment of diagnosis of early/mild SSc according to the Very Early Diagnosis of Systemic Sclerosis (VEDOSS) criteria^73^, or established disease according to the 2013 American College of Rheumatology//European league against rheumatism (ACR/EULAR) classification criteria^74^,
2. Presence of ILD on HRCT at first (baseline) visit as determined by a senior radiologist, and
3. Availability of a baseline HRCT scan with the following settings:
  a. Slice thickness between 0.6 and 3 mm,
  b. One of the following lung kernels available (B60f, B70f, Bl64d, LUNG),
  c. Filtered-back projection as reconstruction algorithm, and
  d. CT image acquired in full inspiration.

For each patient, baseline demographic and clinical parameters, including age, sex, SSc disease duration and subset, extent of skin involvement, autoantibody status, presence of pulmonary hypertension according to right heart catheterization or echocardiography as judged by the local investigators, and pulmonary function test (PFT) parameters were retrieved from the local patients’ records. The recorded PFT parameters (expressed as % predicted values) included forced vital capacity (FVC), forced expiratory volume in 1 second (FEV1), and diffusing capacity for carbon monoxide (DLCO). Data from the 6-minute walk test (6-MWT), including walk distance, oxygen saturation (% SpO2) before and after the test, and Borg scale of perceived exertion (Borg CR-10)^75^, were only available for the derivation cohort. Detailed information about the patients’ demographics and clinical characteristics at baseline for both study cohorts are provided in Supplementary Table 1.

The follow-up period was defined as the time interval between baseline visit and the last available follow-up visit for every patient. The mean follow-up time for the derivation cohort was 66.1 (± 30.1) months and 43.9 (± 30.9) months for the external validation cohort. All outcome events occurring in this period were considered in this study. As clinical outcomes for SSc-ILD we selected 1) progression-free survival and 2) overall survival, which were defined as the time from the date of the HRCT to the date of first occurrence of ILD progression or all-cause death, respectively. Primary endpoint for progression-free survival was the progression of ILD defined as a relative decline in FVC% predicted from baseline to follow-up of ≥ 15%. As a secondary and exploratory endpoint, we used a recently proposed FVC-DLCO composite index, in which progression is defined as either a relative decrease in FVC% predicted of ≥15%, or a relative decline in FVC% predicted of ≥10% combined with DLCO% predicted of ≥15%^45^.

The study was approved by the local ethics committees (approval numbers: pre-BASEC-EK-839 (KEK-no.-2016-01515), KEK-ZH-no. 2010-158/5, BASEC-no. 2018-02165) and written informed consent was obtained from every patient.

### HRCT Image Acquisition and Visual CT Analysis

The settings used for the acquisition of HRCT images are summarized in Supplementary Table 6. All HRCT images were assessed for the presence of characteristic visual features of ILD, including ground glass opacification (GGO), reticular changes, traction bronchiectasis, emphysema, and honeycombing. In addition, the radiological subtype (usual interstitial pneumonia (UIP), nonspecific interstitial pneumonia (NSIP), or diffuse interstitial pneumonia (DIP)) was determined. Further, the extent of lung fibrosis on HRCT, defined as presence of reticular changes and/or honeycombing, was categorized as either <20% or ≥ 20% in relation to the total lung volume. All visual analyses were performed by a senior radiologist (T.F.) using a standard picture archiving and communication system workstation (Impax, Version 6.5.5.1033; Agfa-Gevaert, Mortsel, Belgium) and a high definition liquid crystal display monitor (BARCO; Medical Imaging Systems, Kortrijk, Belgium).

### CT Segmentation and Extraction of Radiomic Features

The left and right lung lobes of each patient were semi-automatically segmented by 2 readers (J.S., M.B.) using the “region grow” function (lower threshold -950 HU, upper threshold: -300 HU) of MIM software (version 6.9.2, MIM Software Inc., Cleveland, Ohio, United States). Manual corrections were applied when computationally defined tissue borders did not coincide with the actual lung borders. In addition, pulmonary hilar vessels and atelectatic lung areas were carefully excluded from the regions of interest.

Radiomic analysis was performed on merged structures of both lung lobes using the in-house developed software Z-Rad based on Python programming language 2.7. For radiomics analysis, CT images were resized to isotropic voxels of 2.75 mm and discretized to a fixed bin size of 50 HU. In total, 1,386 radiomic features were calculated per lung (HU limits: -1000 HU to 200 HU), corresponding to the following radiomic feature classes:

1. Intensity or histogram features (*n* = 17),
2. Texture features (*n* = 137) of the *Gray Level Co-occurrence Matrix* (*n* = 52; GLCM), the *Neighborhood Gray Tone Difference Matrix* (*n* = 5; NGTDM), the *Gray Level Run Length Matrix* (*n* = 32); GLRLM), the *Gray Level Size Zone Matrix* (*n* = 16; GLSZM), the *Gray Level Distance Matrix* (*n* = 16; GLDZM) and the *Neighboring Gray Level Dependence Matrix* (*n* = 16; NGLDM), and
3. Wavelet features (*n* = 1,232).

The first class of radiomic features relates to the histogram or distribution of voxel intensities using first-order statistics (e.g. mean, standard deviation, skewness and kurtosis) and as such quantifies tissue intensity characteristics. The second category including texture features describes the intra-tissue heterogeneity by calculating the statistical, spatial inter-relationship between neighboring voxel intensities ^41^. The third group of features, the wavelet features, calculates the intensity and texture features after wavelet decompositions of the original image using eight different coiflet filters (high-pass to low-pass filters) thereby focusing the features on different frequency ranges^6^.

A list of all radiomic features is provided in Supplementary File 1. Radiomic feature definitions were based on the Imaging Biomarker Standardization Initiative report by Zwanenburg *et al*.^*61*^.

### Assessment of Radiomic Feature Stability

Intraclass correlation (ICC) analysis was performed to assess the stability of radiomic features against intra- and inter-operator variability in the semi-automated segmentation process. For inter-operator ICC analysis, three examiners (J.S., M.B., C.B.), and for intra-operator ICC analysis one examiner (J.S.) twice, independently contoured 15 randomly selected SSc patients from the derivation (Zurich) cohort, and radiomic features were extracted from the multiple delineation structures. The ICC coefficient for every radiomic feature was quantified using two-way mixed effect models and applying the “consistency” method (ICC(3,1)) according to ^76^ using “irr” package of R. Only features with good reproducibility defined as ICC ≥ 0.75^44^ were considered in further analyses.

### Unsupervised Clustering

To identify groups of patients with similar radiomic feature patterns, unsupervised clustering was performed. After confirmation of data clusterability by visual assessment of cluster tendency (VAT) and calculation of the Hopkin’s statistic *H* (with *H* > 0.5 indicating clusterability)^77^, *k*-Means clustering algorithm^78^ was applied to the z-scored radiomic data. Only robust radiomic features (ICC ≥ 0.75) entered the cluster analyses. The optimal number of clusters was determined by varying the number of *k-*clusters between 2 and 10, and selecting the optimal *k* with respect to best visual separation and stability as determined by Jaccard bootstrapping (*n* = 1,000 iterations).

### Building a Quantitative Radiomic Risk Score for SSc-ILD

The Zurich cohort was used as a derivation cohort to build and train the radiomic risk score for ILD progression. Patients with no survival data were excluded from the analysis, resulting in a final dataset of 75 patients. For score building, we adapted a recently described approach by Lu et al^8^ for *z*-scored, radiomic features. Following Lu and colleagues, we selected radiomic features in two steps: 1) Cox regression, and 2) penalized LASSO regression using “cox” family with 10-fold cross validation. In the first step, we applied univariate Cox regression per radiomic feature considering only features with FDR of *p* < 0.005. Features selected in step 1) underwent further reduction by LASSO. Only features with non-zero coefficients were retained. Since limited by the modest sample size of the derivation cohort, we did not perform weighting of score features according to the coefficients from LASSO regression and assigned the same importance to each feature by dividing each standardized feature by the total number of features *j* and constructed the final radiomic score from those features as follows: 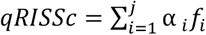 with 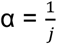 being the feature weightand *f* being the values of *z*-transformed radiomic features.

After having selected features in steps 1) and 2) we searched for the significant cut-off value for Cox regression by applying the “cox” function from the “cutoff” package of R. Due to the modest sample size, we searched for two groups, i.e. “low” and “high” risk patients’ composed of at least 25% of subjects for the minority group. From the proposed pairs of cut-offs, we selected the one that was significant after correction for multiple testing. Once a score was built, we fitted a univariate Cox regression model on the external validation cohort (Oslo). We reported the concordance index (C-index) as the general assessment of the quality of the model, the p-value of the whole model, and the hazard ratio (HR) with 95% confidence intervals for the quantitative radiomic risk score. The C-index is equivalent to the area under the curve in ROC analysis and can also be used in Cox regression analysis^46^. Kaplan-Meier plots were used to visualize the Cox regression results.

### Association Analyses with Clinical Baseline Characteristics and Outcome Measures

Association analyses were performed to explore associations of identified patient groups (*k-*Means clusters and risk groups) with clinical baseline parameters and prognostic outcome measures (progression-free survival and overall survival).

Fisher’s exact test was used for comparison of categorical, and Mann-Whitney U for comparison of continuous clinical variables, respectively.

Kaplan Meier curves and univariate Cox regression were performed to compare progression-free survival and overall survival between the identified patients’ groups for an observation period of up to 90 months.

### Association Analyses with Biological Data

To reveal possible associations of the radiomic risk score with the underlying pathophysiology of ILD, correlation analysis with histological, proteomics and quantitative PCR data was performed. Since lung biopsies are only very rarely performed in SSc-ILD and thus matched patient tissue samples have not been available for molecular analyses, we conducted a cross-species correlation approach, using the mouse model of bleomycin-induced lung fibrosis as model system for SSc-ILD. For this animal model, we have recently confirmed the transferability of radiomics signatures between mice and humans^51^.

#### Animal Model of Experimental ILD

To model human SSc-ILD, we applied the well-established preclinical model of bleomycin-induced lung fibrosis as described previously^54,55^. In brief, 30 female, 8-week-old C57BL/6J-rj (Janvier Labs, Le Genest-Saint-Isle, France) were randomized and intratracheally instilled with 2 U/kg bleomycin sulfate (BLM, Baxter 15,000 I.U., pharmacy of the canton Zurich, Switzerland) to induce ILD. For molecular and histological analyses, mice were sacrificed with carbon dioxide and subsequently transcardially perfused with ice-cold phosphate-buffered saline (PBS) solution to remove residual blood. All animal experiments were approved by the cantonal veterinary office (approval number ZH235-2018) and performed in strict compliance with the Swiss law for animal protection.

#### Proteomic Data

For proteomic analyses, frozen left lung lobes (blood-free) collected from PBS-perfused BLM-treated mice were homogenized in 8M urea/100mM Tris (pH 8.0) buffer supplemented with protease inhibitors using the FastPrep system (MP Biomedicals). After reduction and alkylation, and overnight protein precipitation with ice-cold acetone, 10 ug of the cleaned protein mixture were digested into peptides using a two-step digestion protocol (LysC for 2 h at 37 °C followed by Trypsin at room temperature overnight) and then subjected to liquid-chromatography-based tandem mass spectrometric analysis (LC-MS/MS). For LC-MS/MS, mouse samples were randomly allocated to the analysis by loading 800 ng onto a pre-column (C18 PepMap 100, 5 µm, 100 A, 300 µm i.d. × 5 mm length) at a flow rate of 50µL/min with solvent C (0.05% TFA in water/acetonitrile 98:2).

After loading, peptides were eluted in back flush mode onto a home packed analytical Nano-column (Reprosil Pur C18-AQ, 1.9 µm, 120 A, 0.075 mm i.d. × 500 mm length) using an acetonitrile gradient of 5% to 40% solvent B (0.1% Formic Acid in water/acetonitrile 4,9:95) in 180 min at a flow rate of 250 nL/min. The column effluent was directly coupled to a Fusion LUMOS mass spectrometer (Thermo Fisher, Bremen; Germany) via a nano-spray ESI source. Data acquisition was done in data dependent mode with precursor ion scans recorded in the orbitrap with resolution of 120’000 (at m/z=250) parallel to top speed HCD fragment spectra of the most intense precursor ions in the Linear trap for a cycle time of 3 seconds. Mass spectrometry data was processed by MaxQuant software and set parameters are available in Supplementary Table 7. MaxQuant experimental design was such that the two repeated injections were combined, and match between runs allowed between all samples

#### Histological and Immunohistochemical Data

Formalin-fixed paraffin-embedded lung sections (4 µm thick) from all BLM-treated mice were stained with Hematoxylin and Eosin (HE) for the examination of the overall tissue architecture and the presence of cellular infiltrates, and stained with Picrosirius Red (PSR) to visualize collagen deposition using standard protocols. Furthermore, specific immunohistochemical stainings for the pan-leukocyte marker CD45, and the myofibroblast marker alpha-smooth muscle actin (αSMA) were performed as described in ^54,55^. Whole slide images of histological and immunohistological stainings were obtained with the AxioScan.Z1 slide scanner (Zeiss, Feldbach, Switzerland) in bright-field mode using a Plan-Apochromat 20x/0.8 M27 objective. Stainings were automatically quantified on whole slide images using the open-source Orbit Image Analysis software (License: GPLv3; Actelion Pharmaceuticals Ltd) as described in ^79,80^. Furthermore, for histopathological scoring of pulmonary fibrosis, the Ashcroft score ^56^ was applied on PSR stained lung sections by two experienced blinded examiners (J.S., M.B.) as previously described ^50^.

#### Gene Expression Data

Total RNA was isolated from perfused cranial lobes of the right mouse lung with the RNeasy Tissue Mini Kit from Qiagen (Hombrechtikon, Switzerland), reverse-transcribed into complementary DNA, and messenger RNA (mRNA) expressions of inflammatory (*Il6, Mcp1*) and fibrotic (*Col1a1, Col3a1, Fn1*) genes were analyzed by SYBR Green quantitative real-time PCR as described in ^54^. mRNA expression was expressed as ΔCt values (Ct (gene-of-interest) - Ct (reference gene)) with 60S acidic ribosomal protein P0 (*Rplp0*) as a reference gene, with a lower ΔCt indicating higher target gene expression. A list of primers used in this study is provided in Supplementary Table 8.

#### Micro-CT imaging, Radiomics Analysis and Score Calculation in Mice

CT images were acquired in free-breathing mice with prospective respiratory gating on a state-of-the-art micro-CT scanner (Skyscan 1176; Bruker-microCT, Kontich, Belgium) under isoflurane anesthesia. The following scan parameters were used: tube voltage 50 kV, tube current 500 μA, filter AI 0.5 mm, averaging (frames) 3, rotation step 0.7 degrees, sync with event 50 ms, X-ray tube rotation 360 degrees, resolution 35 μm, and slice thickness 35 μm. Images were reconstructed with NRecon reconstruction software (v.1.7.4.6; Bruker) using the built-in filtered-back projection Feldkamp algorithm and applying misalignment compensation, ring artefact reduction, and a beam hardening correction of 10% to the images.

Analogous to the radiomics analysis in patients, mouse lungs were segmented, resized to isotropic voxels (150 μm) and discretized to a fixed bin size of 50 HU, and all 1,386 radiomic features were extracted (HU limits: -1000 HU to 200 HU). For calculation of the quantitative radiomic ILD risk score, as for patients, the respective radiomic features were *z*-transformed and summed up.

#### Correlation Analysis and Pathway Enrichment Analysis

For correlation analysis with major inflammatory and fibrotic markers on tissue level, Spearman’s rank correlation coefficient *rho* was calculated between the quantitative radiomic ILD risk score and the different biological features.

For pathway enrichment analyses, *rho* was calculated between qRISSc and the LFQ intensity value of every protein that was identified in at least 50% of mice in the proteomics analyses, and only proteins with *p* < 0.05 and *rho* ≥ |0.3| entered further analyses. The resulting list of proteins, and their coding genes, were used as input for the pathway analysis using the ‘ClusterProfiler’ package of Bioconductor. Protein names were converted to gene IDs using the UniProt mapping tool (https://www.uniprot.org/uploadlists/). We investigated pathway enrichment searching against “Reactome” and “GO Biological Process” databases and retained results after adjustment (*p* < 0.05).

### Statistical Analyses

All statistical analyses were conducted in R using the following packages: “ggplot2”, “tidyverse”, “ggsci”, “corrplot”, “readxl”, “clusterSim”, “dplyr”, “readxl”, “survival”, “glmnet”, “cutoff”, “survminer”, “cluster”, “fpc”, “factoextra”, “clustvarsel”, “clustertend”. For all analyses, a p-value of ≤ 0.05 was considered statistically significant.

## Data Availability

For data and material requests please contact the corresponding author.

## Acknowledgements

This work was supported by the Forschungskredit PostDoc from University of Zurich (to JS; FK-19-046), the Bangerter Foundation and Swiss Academy of Medical Sciences (to CM), as well as the Gebauer Foundation, Lunge Zürich Foundation, OPO Foundation, and Prof. Max Cloetta Foundation (to BM). Microscopic image recording was performed with equipment maintained by the Center for Microscopy and Image Analysis, University of Zurich. We acknowledge Maria Comazzi (Center of Experimental Rheumatology, University Hospital Zurich, Switzerland) for her technical assistance with the histological analyses.

## Author Contributions

J.S. contributed to the design and conception of the work, was involved in acquisition, analysis and interpretation of radiomics, clinical and molecular data, and wrote the manuscript. M.M. contributed to the design and conception of the work, performed the statistical analysis, and drafted the manuscript. M.B. contributed to the acquisition and analysis of radiomics, clinical and molecular data, and revised the manuscript. H.S.G. was involved in the acquisition and analysis of radiomics data, and revised the manuscript. C.B. contributed to the acquisition and interpretation of radiomics data, and revised the manuscript. C.M. contributed to the acquisition and interpretation of clinical data, and revised the manuscript. S.B. contributed to the acquisition and interpretation of proteomics data, and revised the manuscript. A.U. contributed to the acquisition and interpretation of proteomics data, and revised the manuscript. M.H. contributed to the acquisition and interpretation of proteomics data, and revised the manuscript. O.D. contributed to the analysis and interpretation of clinical and molecular data, and revised the manuscript. M.G. contributed to the analysis and interpretation of radiomics and clinical data, and revised the manuscript. H.F. contributed to the acquisition and interpretation of clinical data, and revised the manuscript. A.M.H.V. contributed to the acquisition and interpretation of clinical data, and revised the manuscript. C.T.N. contributed to the statistical analysis and interpretation of data, and revised the manuscript T.F. contributed to the acquisition, analysis and interpretation of radiomics data, and revised the manuscript. S.T. contributed to the acquisition, analysis and interpretation of radiomics data and revised the manuscript. B.M. was central for the design and conception of the work, was involved in acquisition, analysis and interpretation of radiomics, clinical and molecular data, and wrote the manuscript.

## Competing Interests

J.S., M.M., H.S.G, M.B., C.M., C.B., M.G., S.B., A.U., M.H., C.T.N., T.F, S.T. have no competing interests to declare. O.D. had consultancy relationship and/or has received research funding from Abbvie, Actelion, Acceleron Pharma, Amgen, AnaMar, Baecon Discovery, Blade Therapeutics, Bayer, Boehringer Ingelheim, Catenion, Competitive Drug Development International Ltd, CSL Behring, ChemomAb, Curzion Pharmaceuticals, Ergonex, Galapagos NV, Glenmark Pharmaceuticals, GSK, Inventiva, Italfarmaco, iQone, iQvia, Lilly, medac, Medscape, Mitsubishi Tanabe Pharma, MSD, Novartis, Pfizer, Roche, Sanofi, Target Bio Science and UCB in the area of potential treatments of scleroderma and its complications. In addition, O.D. has a patent mir-29 for the treatment of systemic sclerosis issued (US8247389, EP2331143). H.F. received travel bursaries from Actelion and GSK, and remuneration from Bayer. A.M.H.V had grant/research support from Boehringer-Ingelheim and received speaker and personal fees from Boehringer-Ingelheim, Roche, Actelion, and Bayer. B. M. had grant/research support from AbbVie, Protagen, Novartis Biomedical Research as well as congress support from Pfizer, Roche, Actelion, and MSD. In addition, B.M. has a patent mir-29 for the treatment of systemic sclerosis registered (US8247389, EP2331143).

## Supplementary Figures

**Supplementary Figure 1:**
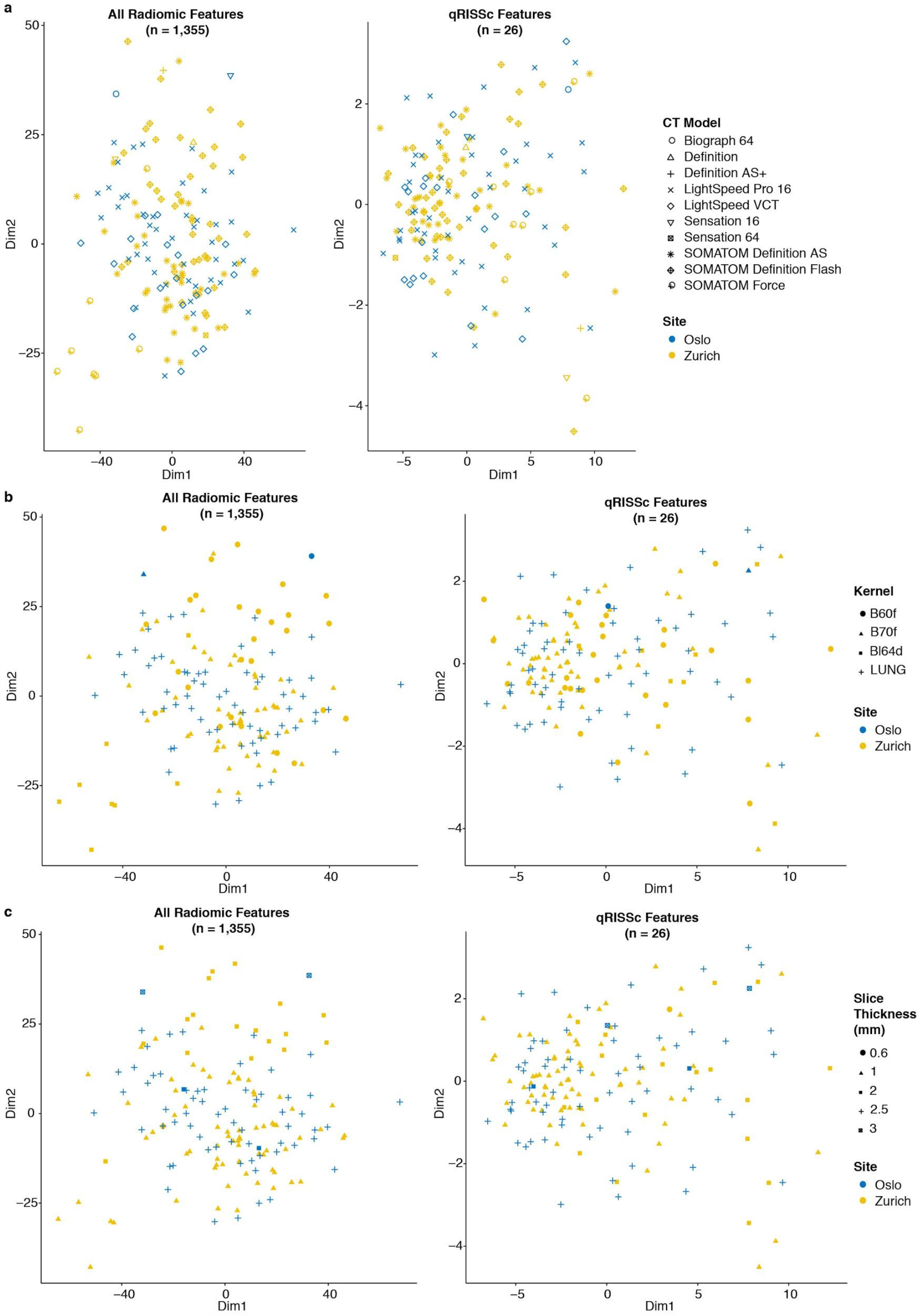
Impact of different CT image acquisition and reconstruction settings on radiomic feature values and qRISSc. Multidimensional scaling of *z*-transformed radiomic profiles of all robust radiomic features (left panel) or only qRISSc features (right panel) combined for all SSc-ILD patients from the Zurich (*n* = 90) and Oslo cohort (*n* = 66) for (**a**) the different CT scanner types, (**b**) different lung reconstruction kernels, and (**c**) different slice thicknesses.

## Supplementary Tables

**Supplementary Table 1:** Summary of patients’ demographics and clinical baseline characteristics for the two study cohorts. Continuous variables are described as median ± interquartile range and categorical variables are presented as absolute counts with relative frequencies (percent). *Abbreviations: UIP = usual interstitial pneumonia, NSIP = nonspecific interstitial pneumonia, DIP = diffuse interstitial pneumonia, PAPsys = systolic pulmonary artery pressure, FVC = forced vital capacity, FEV1 = forced expiratory volume in 1 second, DLCO = diffusing capacity for carbon monoxide, 6-MWT = 6-min walk test*

| Characteristics | Zurich cohort (n=90) | Oslo cohort (n=66) |
| --- | --- | --- |
| **Age (years)** | 57.5 ± 17.8 | 61.0 ± 18.8 |
| **Sex** |  |  |
| Male | 21 (23.3%) | 16 (24.2%) |
| Female | 69 (76.7%) | 50 (75.8%) |
| **SSc disease duration (years)\*** | 5.0 ± 8.2 | 5.3 ± 9.2 |
| **SSc subset (LeRoy 1988)** |  |  |
| Limited cutaneous SSc | 41 (45.6%) | 37 (56.1%) |
| Diffuse cutaneous SSc | 42 (46.7%) | 29 (43.9%) |
| No skin involvement | 7 (7.8%) | 0 (0.0%) |
| **Skin involvement** |  |  |
| Limited cutaneous | 31 (34.4%) | 37 (56.1%) |
| Diffuse cutaneous | 43 (47.8%) | 29 (43.9%) |
| No skin involvement | 9 (10.0%) | 0 (0.0%) |
| Only sclerodactyly | 7 (7.8%) | 0 (0.0%) |
| **Autoantibodies** |  |  |
| Anti-centromere positive | 13 (14.4%) | 7 (10.6%) |
| Anti-topoisomerase I positive | 41 (45.6%) | 24 (36.4%) |
| Anti-RNA polymerase III positive | 7 (7.8%) | 8 (12.1%) |
| Anti-PMScl positive | 18 (20.0%) | 4 (6.1%) |
| **FVC (% predicted)** | 87.5 ± 33.9 | 85.0 ± 36.0 |
| FVC ≥ 70% predicted | 64 (71.1%) | 44 (66.7%) |
| FVC < 70% predicted | 24 (26.7%) | 15 (22.7%) |
| **DLCO (% predicted)** | 66.5 ± 29.4 | 61.0 ± 29.0 |
| **FEV1 (% predicted)** | 88.7 ± 31.2 | 77.0 ± 26.5 |
| **Pulmonary hypertension<sup>†</sup>** | 20 (22.2%) | 6 (9.1%) |
| **PAPsys (mmHg)** | 26.0 ± 10.8 | 15.0 ± 20.0 |
| **6 min walk distance (m)** | 511.0 ± 161.0 | n/a |
| **SpO<sub>2</sub> before 6-MWT (%)** | 96.0 ± 2.0 | n/a |
| **SpO<sub>2</sub> after 6-MWT (%)** | 95.0 ± 7.0 | n/a |
| **Borg scale (unit; range 0-10))** | 3.0 ± 2.0 | n/a |
| **Extent of lung fibrosis on CT** |  |  |
| < 20% | 50 (55.6%) | 30 (45.5%) |
| ≥ 20% | 40 (44.4%) | 36 (54.5%) |
| **Ground glass opacification** | 33 (36.7%) | 42 (63.6%) |
| **Reticular changes** | 86 (95.6%) | 51 (77.3%) |
| **Tractions** | 49 (54.4%) | 27 (40.9%) |
| **Honeycombing** | 23 (25.6%) | 16 (24.2%) |
| **Bullae** | 3 (3.3%) | 4 (6.1%) |
| **Radiological subtype** |  |  |
| NSIP | 76 (84.4%) | 33 (50.0%) |
| UIP | 12 (13.3%) | 28 (42.4%) |
| DIP | 1 (1.1%) | 0 (0.0%) |
| Unclassifiable | 1 (1.1%) | 5 (7.6%) |
| **Immunomodulatory therapy<sup>§</sup>** | 51 (56.7%) | 28 (42.4%) |

\*Disease duration of SSc was calculated as the difference between the date of baseline CT and the date of manifestation of the first non-Raynaud's symptom.
<sup>†</sup>Pulmonary hypertension was assessed by echocardiography or right heart catheterization.
<sup>§</sup>Immunomodulatory therapy included prednisone, methotrexate, rituximab, cyclophosphamide, mycophenolate mofetil, hydroxychloroquine, tocilizumab, imatinib, azathioprine, adalimumab, leflunomide, cyclosporine.

**Supplementary Table 2:** Associations of the identified patients’ clusters based on their radiomic profile with clinical baseline parameters for the Zurich cohort. Continuous variables are described as median ± interquartile range and categorical variables are presented as absolute values with relative frequencies (percent). P-values of univariate comparisons of baseline characteristics between the two clusters are shown. Fisher’s exact test was used for comparison of categorical, and Mann-Whitney U for comparison of continuous variables, respectively. *Abbreviations: UIP = usual interstitial pneumonia, NSIP = nonspecific interstitial pneumonia, DIP = diffuse interstitial pneumonia, PAPsys = systolic pulmonary artery pressure, FVC = forced vital capacity, FEV1 = forced expiratory volume in 1 second, DLCO = diffusing capacity for carbon monoxide, 6-MWT = 6-min walk test*

| Characteristics | Cluster 1 (n=59) | Cluster 2 (n=31) | p value |
| --- | --- | --- | --- |
| **Age (years)** | 58.0 ± 17.0 | 57.0 ± 16.9 | 0.69 |
| **Sex**<br>Male<br>Female | 14 (23.7%)<br>45 (76.3%) | 7 (22.6%)<br>24 (77.4%) | 1 |
| **SSc disease duration (years)\*** | 5.0 ± 8.0 | 4.3 ± 8.3 | 0.51 |
| **SSc subset (LeRoy 1988)**<br>Limited cutaneous SSc<br>Diffuse cutaneous SSc<br>No skin involvement | 30 (50.8%)<br>26 (44.1%)<br>3 (5.1%) | 11 (35.5%)<br>16 (51.6%)<br>4 (12.9%) | 0.23 |
| **Skin involvement**<br>Limited cutaneous<br>Diffuse cutaneous<br>No skin involvement<br>Only sclerodactyly | 20 (33.9%)<br>27 (45.8%)<br>5 (8.5%)<br>7 (11.9%) | 11 (35.5%)<br>16 (51.6%)<br>4 (12.9%)<br>0 (0.0%) | 0.22 |
| **Autoantibodies**<br>Anti-centromere positive<br>Anti-topoisomerase I positive<br>Anti-RNA polymerase III positive<br>Anti-PMScl positive | 10 (16.9%)<br>28 (47.5%)<br>4 (6.8%)<br>14 (23.7%) | 3 (9.7%)<br>13 (41.9%)<br>3 (9.7%)<br>4 (12.9%) | 0.53<br>0.66<br>0.69<br>0.28 |
| **FVC (% predicted)**<br>FVC ≥ 70% predicted<br>FVC < 70% predicted | 97.0 ± 26.0<br>54 (91.5%)<br>4 (6.8%) | 65.5 ± 22.2<br>10 (32.3%)<br>20 (64.5%) | < 0.001 |
| **DLCO (% predicted)** | 75.0 ± 24.0 | 48.0 ± 25.5 | < 0.001 |
| **FEV1 (% predicted)** | 95.8 ± 19.0 | 65.5 ± 25.5 | < 0.001 |
| **Pulmonary hypertension<sup>†</sup>** | 7 (1.9%) | 13 (41.9%) | 0.001 |
| **PAPsys (mmHg)** | 24.0 ± 7.0 | 32.0 ± 18.0 | < 0.001 |
| **6 min walk distance (m)** | 543.5 ± 109.2 | 407.0 ± 173.0 | < 0.001 |
| **SpO<sub>2</sub> before 6-MWT (%)** | 97.0 ± 1.2 | 96.0 ± 3.0 | 0.01 |
| **SpO<sub>2</sub> after 6-MWT (%)** | 96.0 ± 3.0 | 88.5 ± 9.8 | < 0.001 |
| **Borg scale (unit; range 0-10)** | 2.0 ± 2.0 | 4.0 ± 3.0 | < 0.001 |
| **Extent of lung fibrosis on CT**<br>< 20%<br>≥ 20% | 40 (67.8%)<br>19 (32.2%) | 10 (32.3%)<br>21 (67.7%) | 0.002 |
| **Ground glass opacification** | 18 (30.5%) | 15 (48.4%) | 0.11 |
| **Reticular changes** | 58 (98.3%) | 28 (90.3%) | 0.26 |
| **Tractions** | 30 (50.8%) | 19 (61.3%) | 0.37 |
| **Honeycombing** | 9 (15.3%) | 14 (45.2%) | **0.004** |
| **Bullae** | 1 (1.7%) | 2 (6.5%) | 0.24 |
| **Radiological subtype** |  |  |  |
| NSIP | 54 (91.5%) | 22 (71.0%) | **0.01** |
| UIP | 4 (6.8%) | 8 (25.8%) |  |
| DIP | 0 (0.0%) | 1 (3.2%) |  |
| Unclassifiable | 1 (1.7%) | 0 (0.0%) |  |
| **Immunomodulatory therapy<sup>§</sup>** | 29 (49.2%) | 22 (71.0%) | 0.07 |

\*Disease duration of SSc was calculated as the difference between the date of baseline CT and the date of manifestation of the first non-Raynaud's symptom.
<sup>†</sup>Pulmonary hypertension was assessed by echocardiography or right heart catheterization.
<sup>§</sup>Immunomodulatory therapy included prednisone, methotrexate, rituximab, cyclophosphamide, mycophenolate mofetil, hydroxychloroquine, tocilizumab, imatinib, azathioprine, adalimumab, leflunomid, cyclosporine.

**Supplementary Table 3.**
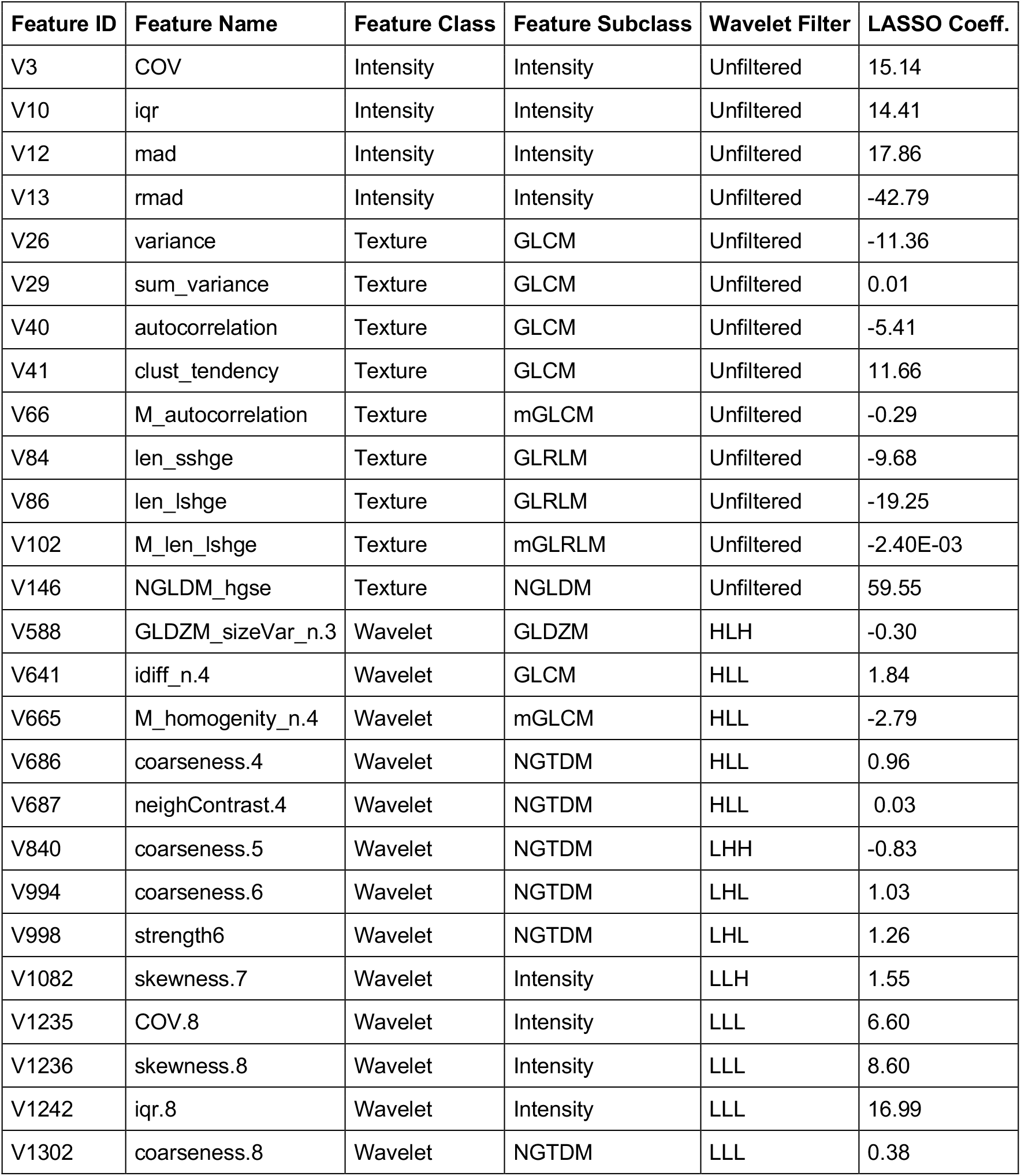
Radiomic features used to construct the quantitative radiomic risk score for SSc-ILD (qRISSc). A full list of all radiomics features including standardized feature names is provided in Supplementary File 1. *Abbreviations: GLCM = Gray Level Co-occurrence Matrix, NGTDM = Neighborhood Gray Tone Difference Matrix, GLRLM = Gray Level Run Length Matrix, GLDZM = Gray Level Distance Matrix and NGLDM = Neighboring Gray Level Dependence Matrix*

| Feature ID | Feature Name | Feature Class | Feature Subclass | Wavelet Filter | LASSO Coeff. |
| --- | --- | --- | --- | --- | --- |
| V3 | COV | Intensity | Intensity | Unfiltered | 15.14 |
| V10 | iqr | Intensity | Intensity | Unfiltered | 14.41 |
| V12 | mad | Intensity | Intensity | Unfiltered | 17.86 |
| V13 | rmad | Intensity | Intensity | Unfiltered | -42.79 |
| V26 | variance | Texture | GLCM | Unfiltered | -11.36 |
| V29 | sum_variance | Texture | GLCM | Unfiltered | 0.01 |
| V40 | autocorrelation | Texture | GLCM | Unfiltered | -5.41 |
| V41 | clust_tendency | Texture | GLCM | Unfiltered | 11.66 |
| V66 | M_autocorrelation | Texture | mGLCM | Unfiltered | -0.29 |
| V84 | len_sshge | Texture | GLRLM | Unfiltered | -9.68 |
| V86 | len_lshge | Texture | GLRLM | Unfiltered | -19.25 |
| V102 | M_len_lshge | Texture | mGLRLM | Unfiltered | -2.40E-03 |
| V146 | NGLDM_hgse | Texture | NGLDM | Unfiltered | 59.55 |
| V588 | GLDZM_sizeVar_n.3 | Wavelet | GLDZM | HLH | -0.30 |
| V641 | idiff_n.4 | Wavelet | GLCM | HLL | 1.84 |
| V665 | M_homogeneity_n.4 | Wavelet | mGLCM | HLL | -2.79 |
| V686 | coarseness.4 | Wavelet | NGTDM | HLL | 0.96 |
| V687 | neighContrast.4 | Wavelet | NGTDM | HLL | 0.03 |
| V840 | coarseness.5 | Wavelet | NGTDM | LHH | -0.83 |
| V994 | coarseness.6 | Wavelet | NGTDM | LHL | 1.03 |
| V998 | strength6 | Wavelet | NGTDM | LHL | 1.26 |
| V1082 | skewness.7 | Wavelet | Intensity | LLH | 1.55 |
| V1235 | COV.8 | Wavelet | Intensity | LLL | 6.60 |
| V1236 | skewness.8 | Wavelet | Intensity | LLL | 8.60 |
| V1242 | iqr.8 | Wavelet | Intensity | LLL | 16.99 |
| V1302 | coarseness.8 | Wavelet | NGTDM | LLL | 0.38 |

**Supplementary Table 4:** Associations of the patients’ risk groups based on qRISSc with clinical baseline parameters for the derivation (Zurich) dataset. Continuous variables are described as median ± interquartile range, and categorical variables are presented as absolute values with relative frequencies (percent). P-values of univariate comparisons of baseline characteristics between the two risk groups are shown. Fisher’s exact test was used for comparison of categorical, and Mann-Whitney U for comparison of continuous variables, respectively. *Abbreviations: UIP = usual interstitial pneumonia, NSIP = nonspecific interstitial pneumonia, DIP = diffuse interstitial pneumonia, PAPsys = systolic pulmonary artery pressure, FVC = forced vital capacity, FEV1 = forced expiratory volume in 1 second, DLCO = diffusing capacity for carbon monoxide, 6-MWT = 6-min walk test*

| Characteristics | Low risk (n=54) | High risk (n=21) | p value |
| --- | --- | --- | --- |
| **Age (years)** | 56.5 ± 16.8 | 56.0 ± 18.0 | 0.87 |
| **Sex**<br>Male<br>Female | 14 (25.9%)<br>40 (74.1%) | 5 (23.8%)<br>16 (76.2%) | 1 |
| **SSc disease duration (years)\*** | 4.3 ± 6.6 | 5.0 ± 9.3 | 0.92 |
| **SSc subset (LeRoy 1988)**<br>Limited cutaneous SSc<br>Diffuse cutaneous SSc<br>No skin involvement | 27 (50.0%)<br>23 (42.6%)<br>4 (7.4%) | 9 (42.9%)<br>10 (47.6%)<br>2 (9.5%) | 0.80 |
| **Skin involvement**<br>Limited cutaneous<br>Diffuse cutaneous<br>No skin involvement<br>Only sclerodactyly | 18 (33.3%)<br>24 (44.4%)<br>6 (11.1%)<br>6 (11.1%) | 9 (42.9%)<br>10 (47.6%)<br>2 (9.5%)<br>0 (0.0%) | 0.48 |
| **Autoantibodies**<br>Anti-Centromere positive<br>Anti-Topoisomerase I positive<br>Anti-RNA polymerase III positive<br>Anti-PMScl positive | 12 (22.2%)<br>28 (51.9%)<br>3 (5.6%)<br>12 (22.2%) | 0 (0.0%)<br>9 (42.9%)<br>3 (14.3%)<br>4 (19.0%) | **0.02**<br>0.61<br>0.34<br>1 |
| **FVC (% predicted)**<br>FVC ≥ 70% predicted<br>FVC < 70% predicted | 97.4 ± 28.5<br>48 (88.9%)<br>6 (11.1%) | 65.0 ± 18.0<br>6 (28.6%)<br>15 (71.4%) | **< 0.001** |
| **DLCO (% predicted)** | 74.4 ± 24.2 | 51.0 ± 25.0 | **< 0.001** |
| **FEV1 (% predicted)** | 95.8 ± 21.0 | 65.0 ± 27.0 | **< 0.001** |
| **Pulmonary hypertension<sup>†</sup>** | 3 (5.6%) | 10 (47.6%) | **< 0.001** |
| **PAPsys (mmHg)** | 24.0 ± 6.8 | 31.0 ± 12.5 | **< 0.001** |
| **6 min walk distance (m)** | 543.0 ± 118.0 | 421.0 ± 126.5 | **0.002** |
| **SpO<sub>2</sub> before 6MWT (%)** | 97.0 ± 1.0 | 96.0 ± 3.2 | 0.09 |
| **SpO<sub>2</sub> after 6MWT (%)** | 96.0 ± 3.0 | 85.5 ± 5.2 | **< 0.001** |
| **Borg (unit; range 0-10)** | 2.0 ± 2.0 | 5.5 ± 3.2 | **< 0.001** |
| **Extent of lung fibrosis on CT**<br>< 20%<br>≥ 20% | 40 (74.1%)<br>14 (25.9%) | 5 (23.8%)<br>16 (76.2%) | **< 0.001** |
| **Ground glass opacification** | 17 (31.5%) | 9 (42.9%) | 0.42 |
| **Reticular changes** | 52 (96.3%) | 19 (90.5%) | 1 |
| **Tractions** | 21 (38.9%) | 16 (76.2%) | **0.003** |
| **Honeycombing** | 5 (9.3%) | 11 (52.4%) | **< 0.001** |
| **Bullae** | 2 (3.7%) | 1 (4.8%) | 1 |
| **Radiological subtype** |  |  |  |
| NSIP | 51 (94.4%) | 14 (66.7%) | **0.002** |
| UIP | 2 (3.7%) | 6 (28.6%) |  |
| DIP | 0 (0.0%) | 1 (4.8%) |  |
| Unclassifiable | 1 (1.9%) | 0 (0.0%) |  |
| **Immunomodulatory therapy<sup>§</sup>** | 30 (55.6%) | 14 (66.7%) | 0.44 |

\*Disease duration of SSc was calculated as the difference between the date of baseline CT and the date of manifestation of the first non-Raynaud's symptom.
<sup>†</sup>Pulmonary hypertension was assessed by echocardiography or right heart catheterization.
<sup>§</sup>Immunomodulatory therapy included prednisone, methotrexate, rituximab, cyclophosphamide, mycophenolate mofetil, hydroxychloroquine, tocilizumab, imatinib, azathioprine, adalimumab, leflunomid, cyclosporine.

**Supplementary Table 5:** Associations of the patients’ risk groups based on qRISSc with clinical baseline parameters for the external and independent validation (Oslo) cohort. Continuous variables are described as median ± interquartile range, and categorical variables are presented as absolute values with relative frequencies (percent). P-values of univariate comparisons of baseline characteristics between the two risk groups are shown. Fisher’s exact test was used for comparison of categorical, and Mann-Whitney U for comparison of continuous variables, respectively. *Abbreviations: UIP = usual interstitial pneumonia, NSIP = nonspecific interstitial pneumonia, DIP = diffuse interstitial pneumonia, PAPsys = systolic pulmonary artery pressure, FVC = forced vital capacity, FEV1 = forced expiratory volume in 1 second, DLCO = diffusing capacity for carbon monoxide, 6-MWT = 6-min walk test*

| Characteristics | Low risk (n=47) | High risk (n=19) | p value |
| --- | --- | --- | --- |
| **Age (years)** | 58.0 ± 22.0 | 64.0 ± 19.0 | 0.59 |
| **Sex**<br>Male<br>Female | 12 (25.5%)<br>35 (74.5%) | 4 (21.1%)<br>15 (78.9%) | 1 |
| **Disease duration (years)\*** | 4.3 ± 9.1 | 6.1 ± 9.1 | 0.33 |
| **SSc subset (LeRoy 1988)**<br>Limited cutaneous SSc<br>Diffuse cutaneous SSc<br>No skin involvement | 26 (55.3%)<br>21 (44.7%)<br>0 (0.0%) | 11 (57.9%)<br>8 (42.1%)<br>0 (0.0%) | 1 |
| **Skin involvement**<br>Limited cutaneous<br>Diffuse cutaneous<br>No skin involvement<br>Only sclerodactyly | 26 (55.3%)<br>21 (44.7%)<br>0 (0.0%)<br>0 (0.0%) | 11 (57.9%)<br>8 (42.1%)<br>0 (0.0%)<br>0 (0.0%) | 1 |
| **Autoantibodies**<br>Anti-Centromere positive<br>Anti-Topoisomerase I positive<br>Anti-RNA polymerase III positive<br>Anti-PMScl positive | 5 (10.6%)<br>17 (36.2%)<br>8 (17.0%)<br>3 (6.4%) | 2 (10.5%)<br>7 (36.8%)<br>0 (0.0%)<br>1 (5.3%) | 1<br>1<br>0.05<br>1 |
| **FVC (% predicted)**<br>FVC ≥ 70% predicted<br>FVC < 70% predicted | 92.0 ± 25.5<br>36 (76.6%)<br>6 (12.8%) | 60.0 ± 20.0<br>8 (42.1%)<br>9 (47.4%) | **< 0.001** |
| **DLCO (% predicted)** | 66.0 ± 17.5 | 35.0 ± 20.0 | **< 0.001** |
| **FEV1 (% predicted)** | 82.0 ± 22.0 | 64.0 ± 18.0 | **0.001** |
| **Pulmonary hypertension<sup>†</sup>** | 1 (2.1%) | 5 (26.3%) | **0.01** |
| **PAPsys (mmHg)** | 15.0 ± 20.0 | 35.0 ± 17.5 | 0.13 |
| **6 min walk distance (m)** | n/a | n/a | n/a |
| **SpO<sub>2</sub> before 6MWT (%)** | n/a | n/a | n/a |
| **SpO<sub>2</sub> after 6MWT (%)** | n/a | n/a | n/a |
| **Borg (unit; range 0-10))** | n/a | n/a | n/a |
| **Extent of lung fibrosis on CT**<br>< 20%<br>≥ 20% | 30 (63.8%)<br>17 (36.2%) | 0 (0.0%)<br>19 (100.0%) | **< 0.001** |
| **Ground glass opacification** | 33 (70.2%) | 9 (47.4%) | 0.10 |
| **Reticular changes** | 34 (72.3%) | 17 (89.5%) | 0.20 |
| **Tractions** | 12 (25.5%) | 15 (78.9%) | **< 0.001** |
| **Honeycombing** | 6 (12.8%) | 10 (52.6%) | **< 0.001** |
| **Bullae** | 2 (4.3%) | 2 (10.5%) | 0.57 |
| **Radiological subtype** |  |  |  |
| NSIP | 26 (55.3%) | 7 (36.8%) | 0.16 |
| UIP | 17 (36.2%) | 11 (57.9%) |  |
| DIP | 0 (0.0%) | 0 (0.0%) |  |
| Unclassifiable | 4 (8.5%) | 1 (5.3%) |  |
| **Immunomodulatory therapy<sup>§</sup>** | 16 (34.0%) | 12 (63.2%) | 0.05 |

\*Disease duration of SSc was calculated as the difference between the date of baseline CT and the date of manifestation of the first non-Raynaud's symptom.
<sup>†</sup>Pulmonary hypertension was assessed by echocardiography or right heart catheterization.
<sup>§</sup>Immunomodulatory therapy included prednisone, methotrexate, rituximab, cyclophosphamide, mycophenolate mofetil, hydroxychloroquine, tocilizumab, imatinib, azathioprine, adalimumab, leflunomid, cyclosporine.

**Supplementary Table 6:** Summary of HRCT image acquisition parameters for the two study cohorts. For slice thickness and tube voltage, data are presented as median and range of minimal and maximal values.

| CT parameter | Discovery (Zurich) cohort (n=90) | Validation (Oslo) cohort (n=66) |
| --- | --- | --- |
| Manufacturer* | Siemens | Siemens, GE Medical Systems |
| Acquisition Model | Inspiration (breath hold) | Inspiration (breath hold) |
| Slice thickness (mm) | 1 (range 0.6 - 2) | 2.5 (range 2 - 3) |
| Reconstruction kernels | B60f, B70f, BI64 | B60f, B70f, LUNG |
| Tube voltage (kVP) | 120 (range 80 - 150) | 120 |

\*HRCT scanners included SOMATOM Definition AS, SOMATOM Definition Flash, SOMATOM Force, SOMATOM Sensation 64, SOMATOM Sensation 16, Biograph 64, LightSpeed Pro 16, LightSpeed VCT.

**Supplementary Table 7:**
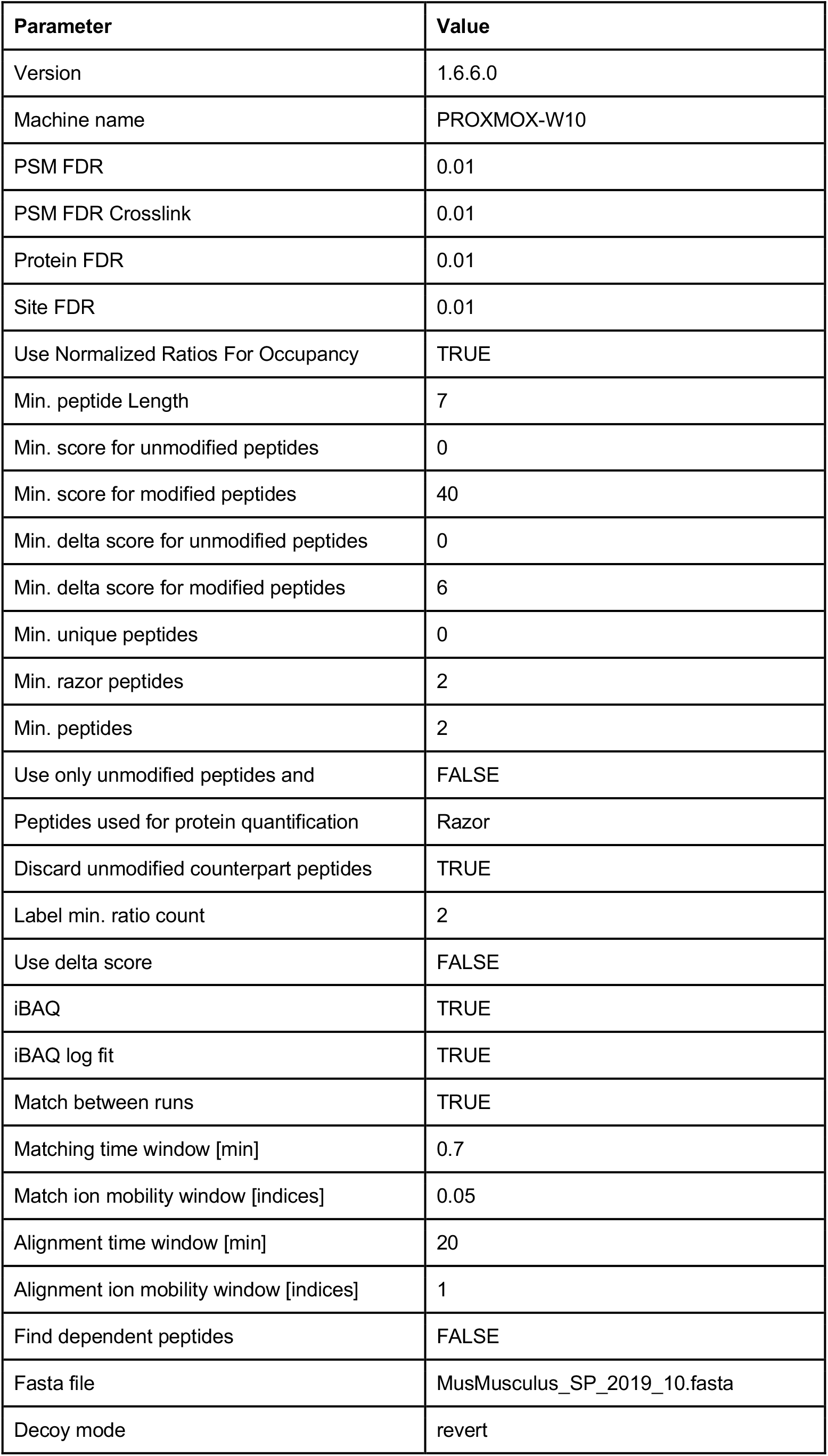
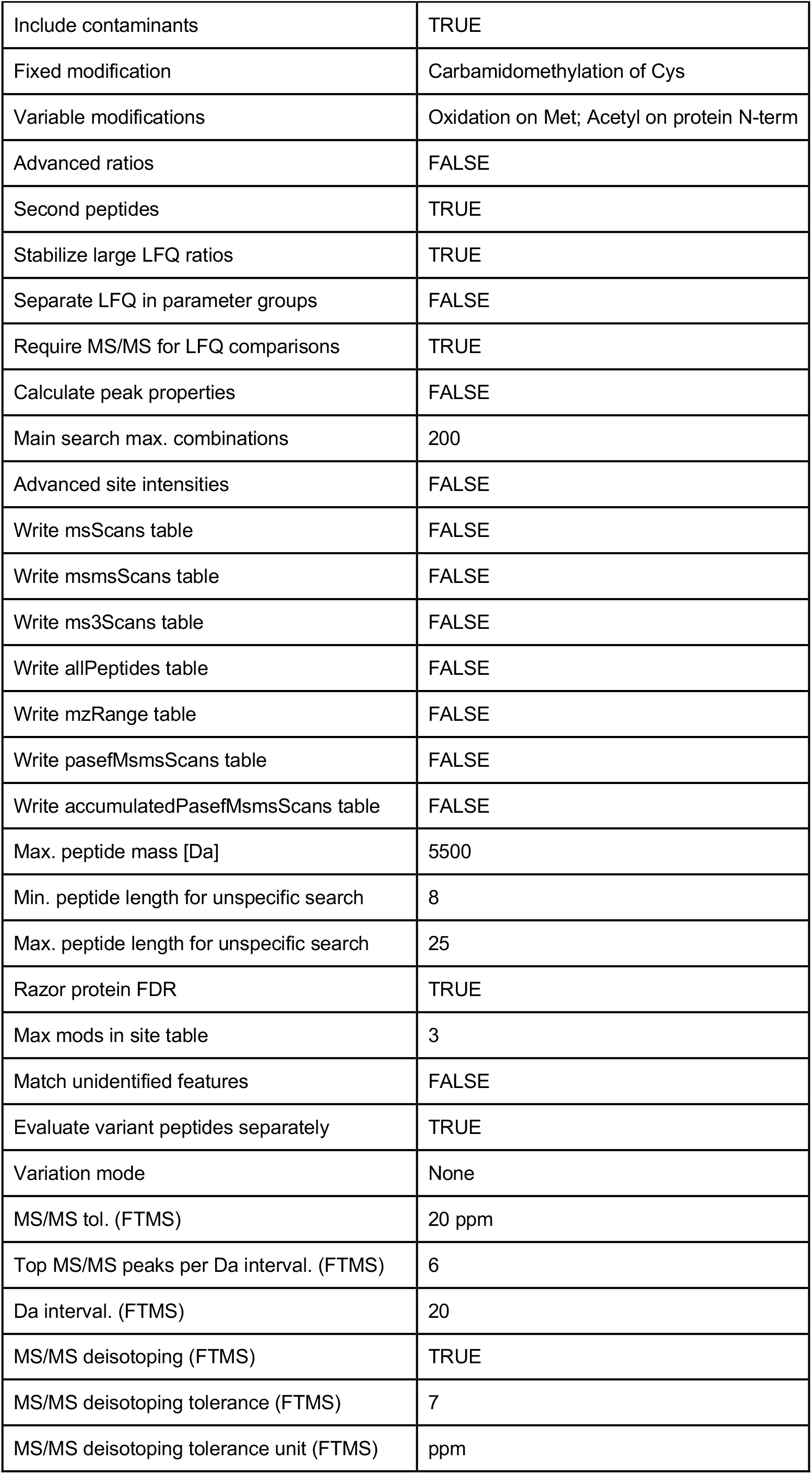
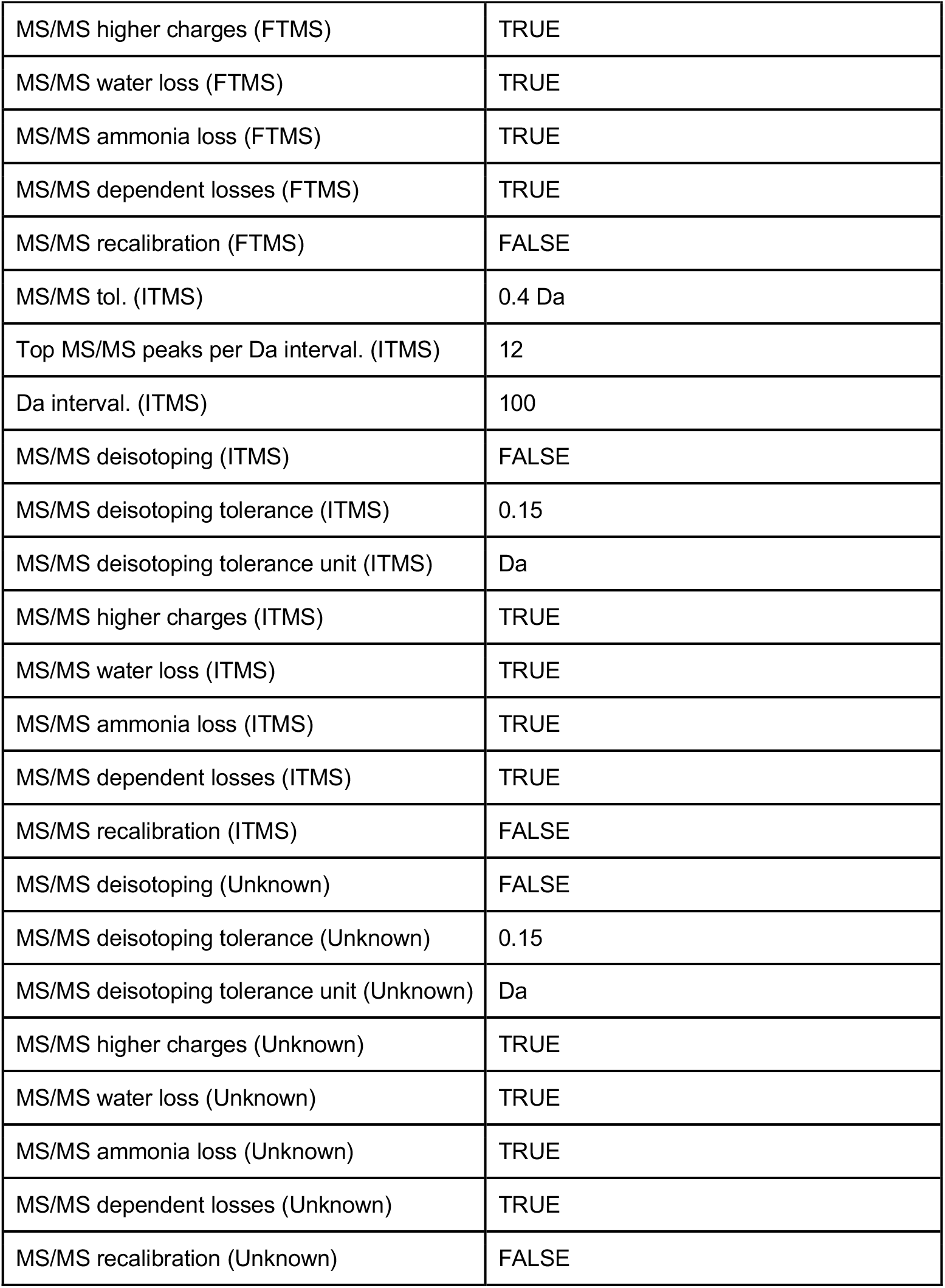
Parameter settings for MaxQuant analysis.

| Parameter | Value |
| --- | --- |
| Version | 1.6.6.0 |
| Machine name | PROXMOX-W10 |
| PSM FDR | 0.01 |
| PSM FDR Crosslink | 0.01 |
| Protein FDR | 0.01 |
| Site FDR | 0.01 |
| Use Normalized Ratios For Occupancy | TRUE |
| Min. peptide Length | 7 |
| Min. score for unmodified peptides | 0 |
| Min. score for modified peptides | 40 |
| Min. delta score for unmodified peptides | 0 |
| Min. delta score for modified peptides | 6 |
| Min. unique peptides | 0 |
| Min. razor peptides | 2 |
| Min. peptides | 2 |
| Use only unmodified peptides and | FALSE |
| Peptides used for protein quantification | Razor |
| Discard unmodified counterpart peptides | TRUE |
| Label min. ratio count | 2 |
| Use delta score | FALSE |
| iBAQ | TRUE |
| iBAQ log fit | TRUE |
| Match between runs | TRUE |
| Matching time window [min] | 0.7 |
| Match ion mobility window [indices] | 0.05 |
| Alignment time window [min] | 20 |
| Alignment ion mobility window [indices] | 1 |
| Find dependent peptides | FALSE |
| Fasta file | MusMusculus_SP_2019_10.fasta |
| Decoy mode | revert |
| Include contaminants | TRUE |
| Fixed modification | Carbamidomethylation of Cys |
| Variable modifications | Oxidation on Met; Acetyl on protein N-term |
| Advanced ratios | FALSE |
| Second peptides | TRUE |
| Stabilize large LFQ ratios | TRUE |
| Separate LFQ in parameter groups | FALSE |
| Require MS/MS for LFQ comparisons | TRUE |
| Calculate peak properties | FALSE |
| Main search max. combinations | 200 |
| Advanced site intensities | FALSE |
| Write msScans table | FALSE |
| Write msmsScans table | FALSE |
| Write ms3Scans table | FALSE |
| Write allPeptides table | FALSE |
| Write mzRange table | FALSE |
| Write pasefMsmsScans table | FALSE |
| Write accumulatedPasefMsmsScans table | FALSE |
| Max. peptide mass [Da] | 5500 |
| Min. peptide length for unspecific search | 8 |
| Max. peptide length for unspecific search | 25 |
| Razor protein FDR | TRUE |
| Max mods in site table | 3 |
| Match unidentified features | FALSE |
| Evaluate variant peptides separately | TRUE |
| Variation mode | None |
| MS/MS tol. (FTMS) | 20 ppm |
| Top MS/MS peaks per Da interval. (FTMS) | 6 |
| Da interval. (FTMS) | 20 |
| MS/MS deisotoping (FTMS) | TRUE |
| MS/MS deisotoping tolerance (FTMS) | 7 |
| MS/MS deisotoping tolerance unit (FTMS) | ppm |
| MS/MS higher charges (FTMS) | TRUE |
| MS/MS water loss (FTMS) | TRUE |
| MS/MS ammonia loss (FTMS) | TRUE |
| MS/MS dependent losses (FTMS) | TRUE |
| MS/MS recalibration (FTMS) | FALSE |
| MS/MS tol. (ITMS) | 0.4 Da |
| Top MS/MS peaks per Da interval. (ITMS) | 12 |
| Da interval. (ITMS) | 100 |
| MS/MS deisotoping (ITMS) | FALSE |
| MS/MS deisotoping tolerance (ITMS) | 0.15 |
| MS/MS deisotoping tolerance unit (ITMS) | Da |
| MS/MS higher charges (ITMS) | TRUE |
| MS/MS water loss (ITMS) | TRUE |
| MS/MS ammonia loss (ITMS) | TRUE |
| MS/MS dependent losses (ITMS) | TRUE |
| MS/MS recalibration (ITMS) | FALSE |
| MS/MS deisotoping (Unknown) | FALSE |
| MS/MS deisotoping tolerance (Unknown) | 0.15 |
| MS/MS deisotoping tolerance unit (Unknown) | Da |
| MS/MS higher charges (Unknown) | TRUE |
| MS/MS water loss (Unknown) | TRUE |
| MS/MS ammonia loss (Unknown) | TRUE |
| MS/MS dependent losses (Unknown) | TRUE |
| MS/MS recalibration (Unknown) | FALSE |

**Supplementary Table 8:**
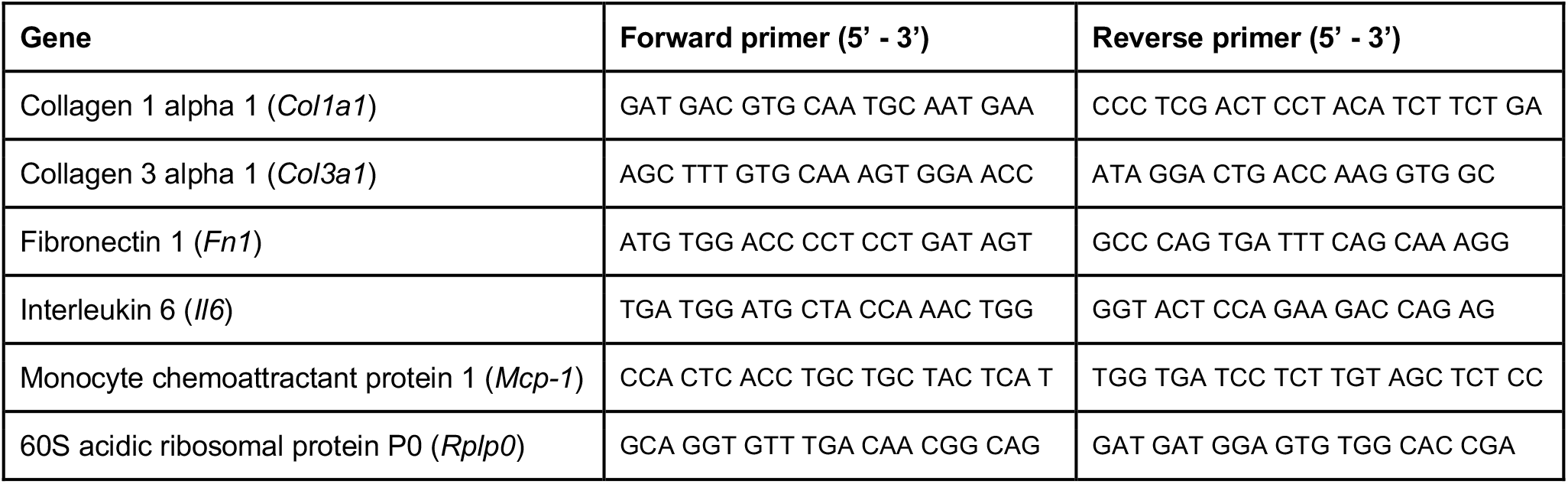
Murine primer sequences used for qRT-PCR.

## Notes

### Funding Statement

This work was supported by the Forschungskredit PostDoc from University of Zurich (to J.S.; FK-19-046), the Bangerter Foundation and Swiss Academy of Medical Sciences (to C.M.), as well as the Gebauer Foundation, Lunge Zurich Foundation, OPO Foundation, and Prof. Max Cloetta Foundation (to B.M.).

